# Technical report: A comprehensive comparison between different quantification versions of Nightingale Health’s ^1^H-NMR metabolomics platform

**DOI:** 10.1101/2023.07.03.23292168

**Authors:** D. Bizzarri, M.J.T. Reinders, M. Beekman, BBMRI-NL, P.E. Slagboom, E.B. van den Akker

## Abstract

^1^H-NMR metabolomics data is increasingly used to track various aspects of health and disease. With the availability of larger data resources and continuously improving learning algorithms Nightingale Health has recently updated the quantification and calibration strategy of their platform to further align their reported analytes with clinical standards. Such updates, however, might influence backward replicability and could hamper comparison of repeated measures in longitudinal studies. Based on data of the BBMRI.nl consortium (>25.000 samples across 28 studies), we compared Nightingale data, as originally released in 2014 and 2016, with a re-quantified version of this data released in 2020, of which both versions were based on the same original NMR spectra. Apart from 2 discontinued, and 23 newly defined analytes, we overall observe a high concordance between quantification versions, with 73 out of 222 (33%) showing a mean correlation > 0.9 across the 28 Dutch cohorts. Nevertheless, five metabolites consistently showed relatively low correlations (R<0.7) between platform versions, namely acetoacetate *(acace),* LDL particle size *(ldl_d)*, saturated fatty acids percentage *(sfa_fa)*, S-HDL-C *(s_hdl_c)* and sphingomyelins (*sm)*. Previously trained multi-analyte scores, such as our previously published health predictors *MetaboAge* or *MetaboHealth*, might be particularly sensitive to platform changes. Whereas the *MetaboHealth* score replicated well between platform versions, the *MetaboAge* score indeed had to be retrained due to discontinued metabolites. Notably, both scores projected on the 2020 re-quantified data did recapitulate the original mortality associations observed in the previous version of the data. Concluding, we urge caution when utilizing data from different quantification versions to avoid mixing analytes capturing different underlying aspects of the NMR spectra, having different units, or simply being discontinued.

## Introduction

Targeted ^1^H-NMR Metabolomics has rapidly gained popularity as a cost-effective and comprehensive method to perform metabolic profiling and risk prediction in large epidemiological studies. Various of such metabolomics-based age predictors were constructed, for example *MetaboAge*, an indicator of several future cardiovascular diseases [1] and *MetaboHealth* that predicts multiple health conditions and all-cause mortality [2]. Thus far, targeted ^1^H-NMR Metabolomics has shown promise to predict COVID hospitalization [3], various disease outcomes [4,5], and a plethora of conventional clinical risk variables [6].

Targeted ^1^H-NMR approaches focus on the analysis of a limited and pre-defined set of analytes, whose associated peaks consistently appear at relatively fixed positions in the overall NMR spectrum of a specific biomaterial and can therefore be robustly quantified. Each of the associated peaks are quantified according to standardized rules and then transformed into absolute quantities with the aid of reference compounds [7]. While each change in the assayed biomaterials or isolation protocols would necessitate a considerable effort to re-calibrate a ^1^H-NMR-based quantification setup, a rigid standardization of both the input material and the laboratory routines would allow for a cost-effective and metabolome profiling on an epidemiological scale.

Nightingale Health Plc is a major commercial supplier of targeted ^1^H-NMR metabolomics data with bench-to-data solutions for human serum, plasma, or urine, for a limited number of metabolic markers. Large consortia like BBMRI.NL [1], FINSK/THL [5], COMETS [8], and more recently UK-Biobank [3] have set out to enrich their population studies with ^1^H-NMR metabolomics profiling and to date have accumulated data in respectively ∼35.000, ∼40.000,

∼46.000, and ∼300.000 samples. Sample handling and processing inevitably varies during and between such large efforts and may introduce variation in the data that could potentially impede replication efforts. In parallel with their metabolomics profiling efforts in UK-Biobank, Nightingale Health updated the way their analytes are quantified to further improve the calibration of 37 of their analytes with clinically measured counterparts. While such updates constitute a further optimization of this biomarker platform, it may also introduce systematic changes with respect to previously assayed studies [9].

Here, we set out to quantify to which extent the most recent updates of the quantification procedure by Nightingale affected the reported analytes, and to what extent this could influence replication of previous findings. We investigated the metabolic-specific correlations in ∼220 features, quantified by Nightingale Health, available for the *same* samples across three different platform versions (2014, 2016 and 2020). We found that, while many analytes present a high degree of correlation between versions, a number of analytes present a moderate to low correlation. In addition, we demonstrate that the effect on multi-analyte scores may differ, and thus ideally would require their renewed validation for each platform update. For example, the *MetaboHealth* score exhibits similar associations with time to death, whereas the metabolomics-based age predictor (*MetaboAge*) could no longer be readily applied due to use of discontinued metabolites yet could be successfully retrained on the new platform version and showed similar associations with disease outcomes.

## Results

All comparisons are done on data gathered within the BBMRI.NL consortium (∼35,000 samples in 28 cohorts, Methods Table 1). Samples were assayed using the Nightingale Health platform in multiple waves of data generation, as indicated with their respective years: 2014 and 2016. After the complete platform update by Nightingale of 2020, BBMRI.NL decided to re-quantify their dataset completely to have metabolomics features comparable to other consortia. It is important to stress that re-quantification consisted of a novel (computational) analyte quantification of the original assays performed in 2014 and 2016; i.e., no new samples were assayed.

**Table 1:** Data and platform versions available in BBMRI.NL

|  | N. Samples | N. Biobanks | Platform version |
| --- | --- | --- | --- |
| *First wave* | 24,994 | 26 | Version 2014 |
| *Second wave* | 9,880 | 10 | Version 2016 |
| *re-quantifications* | 34,015 | 28 | Version 2020 |

**Table 2:**
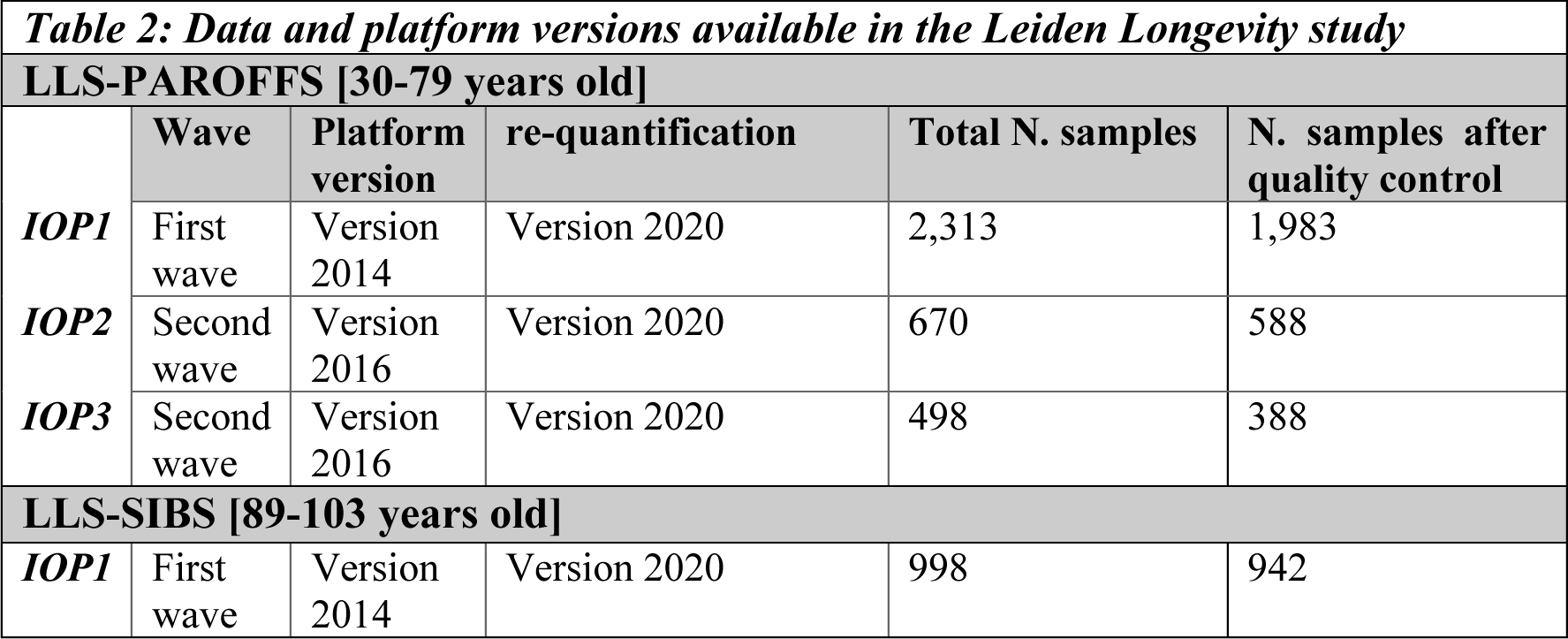
Data and platform versions available in the Leiden Longevity study

### An overview of changes in measured metabolic features

With respect to marker availability, there are new and discontinued reported entities. Notably, the latest version of the platform (2020 version) includes 37 biomarkers which have been CE-approved for diagnostic purposes, i.e., ‘clinically validated’, making the Nightingale platform now not only interesting for epidemiological research, but also suited for use in the clinic [10]. In addition, 25 new variables were added to the pool of metabolic markers now also readily measurable in EDTA plasma (Supplementary Materials). Moreover, the analyte pyruvate (pyr) is featured on the platform again, after being discontinued in 2016. Conversely, analytes showing insufficient replicability were discontinued, either already in the 2016 version (*dag*, *dagtg*, *fallen*, *cla*, *cla_fa*), or from 2020 onward (*hdl2_c* and *hdl3_c*), thus posing potential backward compatibility issues.

Looking more closely to the data, we also note some more subtle changes that nevertheless are helpful to highlight. Compared to older platform versions, the proportion of problematic values decreased in the re-quantified version of the platform, i.e., there are less values that failed to be detected (NaNs), were reported as zero, or were considered outliers (**Figure S1**). In addition, we observe that some markers were reported using different units between, and occasionally within, platform versions. For instance, albumin (alb) changed unit from [signal area] in 2014 to [g/L] in 2020. Particularly interesting are the different ranges of creatinine in the re-quantified measurements (2020 version), which in our case seems to depend on whether the first Nightingale metabolomics quantification was done either in 2014 or in 2016, with reported units in mmol/L and μmol/L respectively (Figure S2). These changes, if unnoticed, can impair replication of the results and application of multi-variate models.

### Correlation analyses of metabolomics measurements between platform versions

First, we evaluated the correlation for each homonymous metabolic measurement across the different Nightingale platform versions within the Leiden Longevity study (LLS); a two-generation cohort containing highly aged individuals (LLS-SIBS) and their offspring with the relative partners (LLS-PAROFFS), with repeated measures over different time-points (IOP1, 2 and 3) (detailed description in *Material and Methods*). Considering same samples of LLS-PAROFFS IOP1, measured the first time in 2014 and re-quantified in 2020 (Figure 1A), we observed that 36 out of the 65 homonymous non-derived analytes (55%), showed a correlation higher than 0.9, with one having a perfect correlation (*glucose*). 24 had a medium correlation (0.7 ≤ R < 0.9), and only five analytes had a correlation lower than 0.7 (*acace, ldl_d*, *sfa_fa*, *s_hdl_c* and *sm*). Some analytes showed a shift in mean values, presumably as a result of a recalibration step, as reflected by a change in levels, e.g. *ldl_d*: first wave [22.99÷25.5 nm] vs. re-quantified [23.4÷24.09 nm], or in units, e.g. *alb:* first wave [0.06÷0.14 signal area] vs. re-quantified [25.6÷62.78 g/l]. Furthermore, also 54 out of the remaining 169 analytes, mostly containing derived measures, showed lower correlations (R<0.7) (Figure S4).

**Figure 1:**
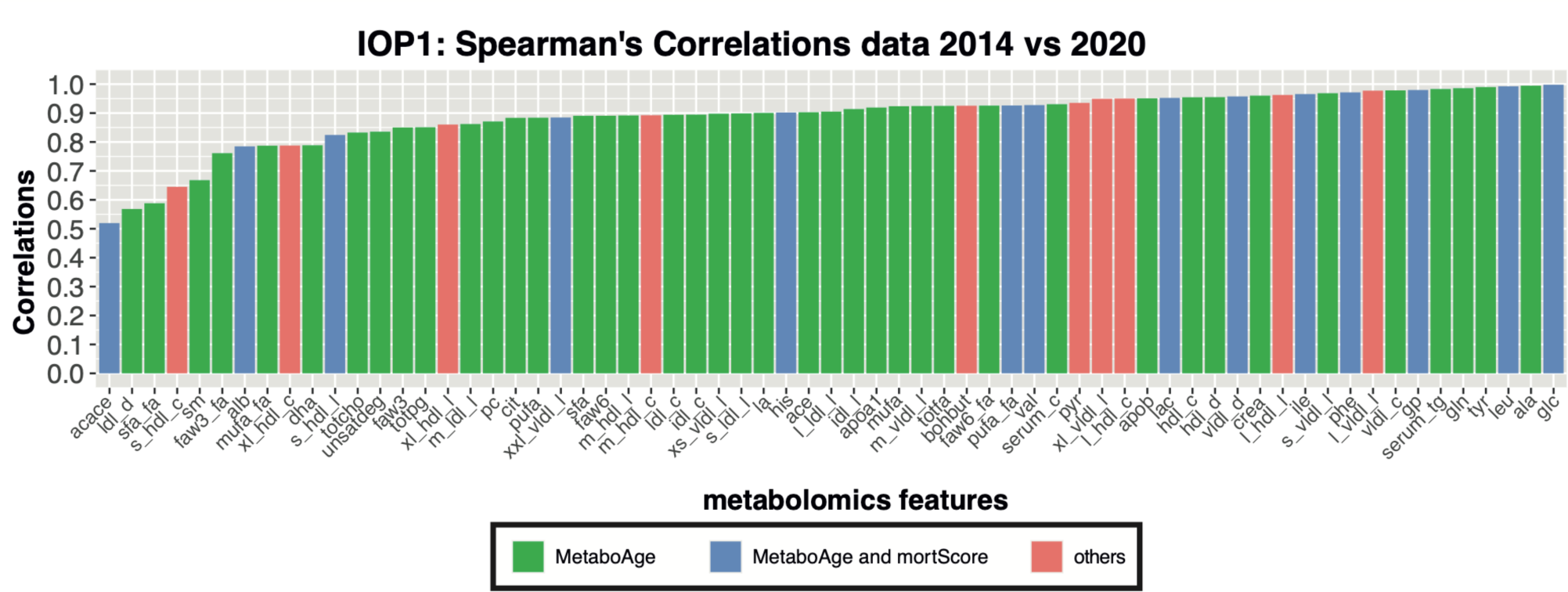
*Evaluation of the metabolic markers before and after re-quantification in LLS-PAROFFS IOP1:* [A] Spearman’s correlations of the homonymous analytes measured in the first wave (2014) with their re-quantified version (2020).

**Figure 2:**
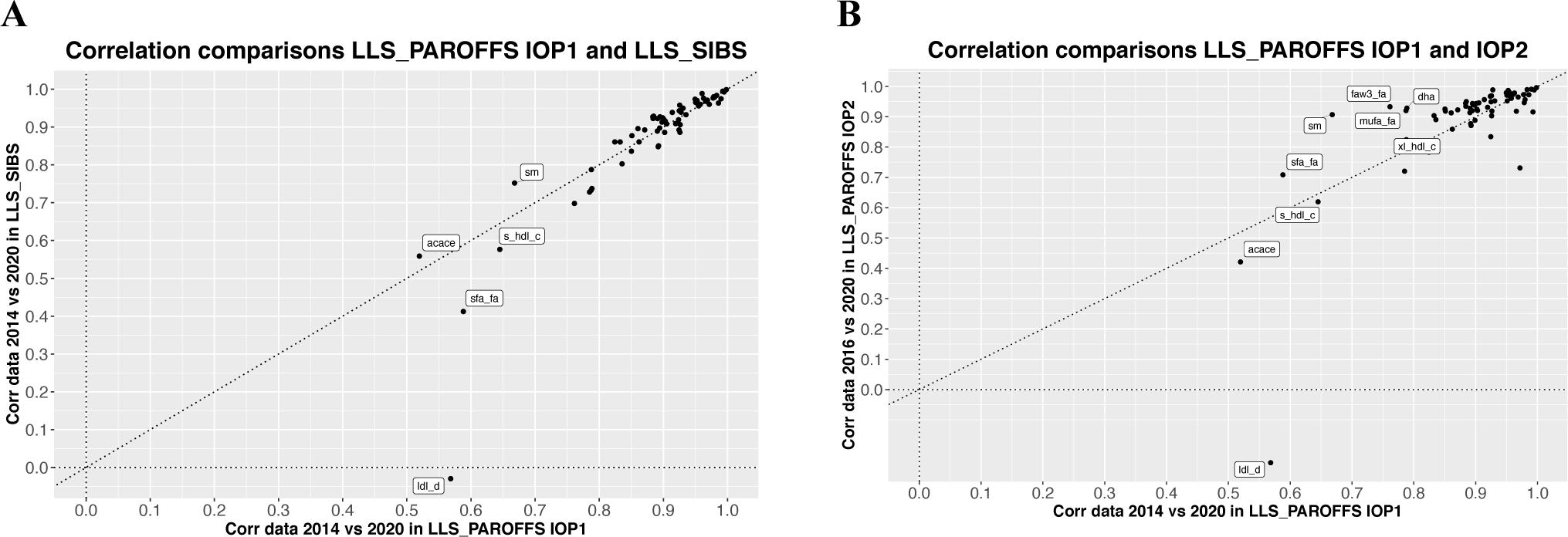
*Comparisons of the correlations of the metabolites before and after re-quantification in different subgroups or platform versions:* Each point of the scatterplots indicate the correlations of each metabolic markers before and after the re-quantifications in [A] LLS-PAROFFS IOP1 (x axis, first measured in 2014) and LLS-SIBS (y axis, first measured in 2014); and [B] LLS-PAROFFS IOP1 (x axis, quantification version 2014) with LLS-PAROFFS IOP2 (y axis, quantification version 2016). Metabolic markers were tagged if they show differences in correlations.

When computing the same correlation analyses comparing LLS_PAROFFS IOP1 (2014 data) with another cohort measured in the first wave, LLS-SIBS (2014 data), or with data of the same cohort of the second wave LLS_PAROFFS IOP2 (2016 data), we observe highly similar trends (Fig2A and S5A). While the majority of analytes show consistently high correlations with their re-quantified counterpart across waves and cohorts, we do observe some notable exceptions. Analytes with a low calibration correlation (R<0.7) in the first data wave (either LLS_PAROFFS, or LLS-SIBS 2014 data) seem to show improvement in the second wave data (either LLS_PAROFFS, 2016 data), except for *ldl_d*. Considering that we find similar results also in LLS-PAROFFS IOP3 (Figure S5C-D), a second round of repeated measures quantified with the Nightingale platform 2016, we concluded that this latter platform version is more similar to the re-quantified data as compared to 2014 version.

To investigate how the correlations of metabolomic features between the different Nightingale platform version behave over different cohorts, we examined these on the whole BBMRI.NL dataset comprising 28 cohorts (***Figure 3***). Observed correlations vary between −0.5 (generally for derived analytes, such as ratios or percentages) and perfect positive correlation (*glucose*). The lower correlations were not due to a lower variance in the markers (Figure S5). Even though there are some cohorts that show generally lower correlations for all the analytes (e.g., BIOMARCS, or STEMI-GIPS), the other cohorts show consistent correlations for the different analytes. 73 analytes had a mean correlation above 0.9 across all BBMRI.NL biobanks (***Figure S6C, Table 5)***. 33 and 8 of these markers overlap with the 65 (51%) and 14 (57%) analytes that were used to construct the *MetaboAge* and *MetaboHealth* score respectively (Figure S6D).

**Figure 3:**
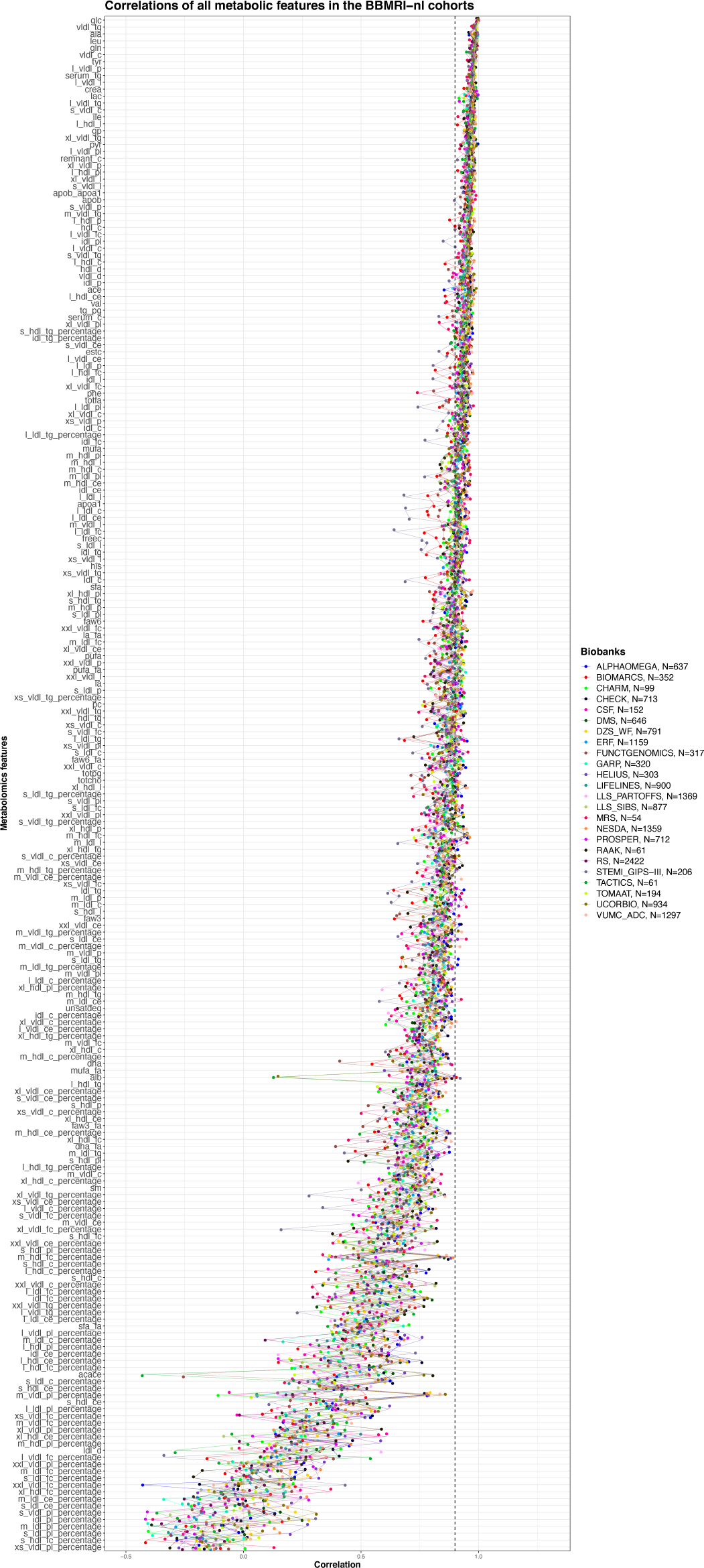
*Correlations of all the metabolomics features before and after re-quantification in all the BBMRI.NL cohorts in data 2014 vs data 2020:* Each dot represents the correlation of a metabolomic feature with itself, colored based on the cohorts included in BBMRI.NL. Dots of the same cohort are connected with a line to highlight outlier patterns. A horizontal dotted line indicates a correlation of 0.9.

### The clinically validated biomarkers show similar correlation, but improved calibration with respect to previous quantification

The latest Nightingale metabolomics platform contains 37 analytes approved by the European community for diagnostics [10]. This is particularly interesting for Consortia like BBMRI.NL, as it allows for an efficient quantification of various routinely assessed clinical biomarkers in one single platform. For this purpose, we evaluated to what extent previously measured clinical variables within BBMRI.NL align with their corresponding analytes on the Nightingale platform. Four of the 37 clinical biomarkers (HDL-cholesterol, LDL-cholesterol, triglycerides, and total cholesterol) were available in 13 of the 28 cohorts (14,995 samples, Figure 4) and showed a medium to high correlation in most of the cohorts, apart for BIOMARCS, PROSPER, and UCORBIO [mean R= 0.6]. While different Nightingale versions generally showed very similar correlations with their clinical chemistry counterparts, notable differences are observed when considering the Median Absolute Distance (MAD). For the 2020 version, we observe an improved concordance between clinically measured biomarkers and their Nightingale counterpart, particularly for LDL-cholesterol and total cholesterol.

**Figure 4:**
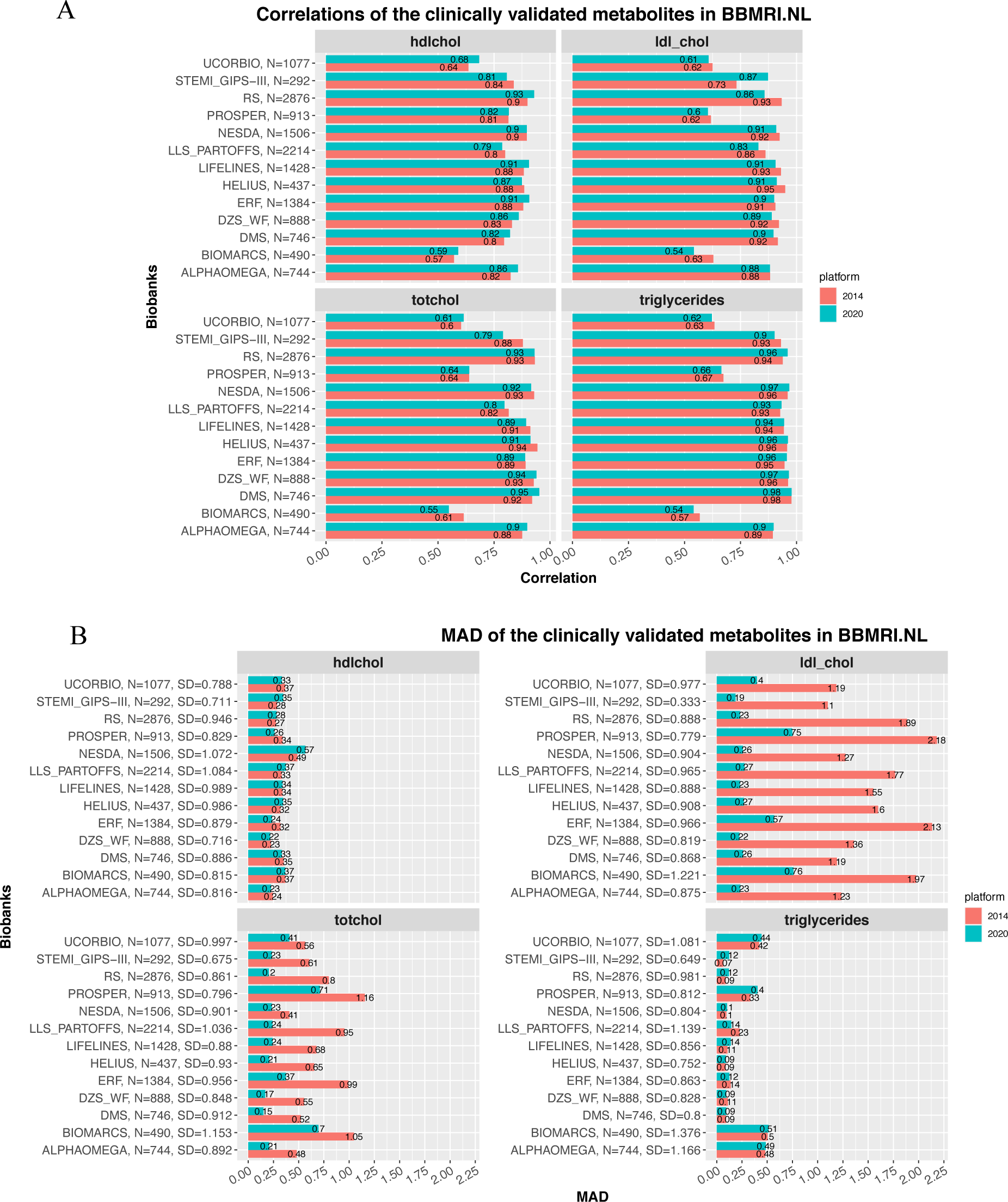
*Correlations of Nightingale metabolomics markers, measured in 2014 and 202, with the clinically measured values in BBMRI.NL:* Bar-plots of the [A] spearman’s correlations and [B] the Median Absolute Distance (MAD) of the *hdl cholesterol*, *ldl cholesterol* (calculated with the Friedewald equation), total cholesterol and triglycerides calculated with clinical chemistry, with their corresponding values in the Nightingale assay (*hdl_c*, *ldl_c/clinical_ldl_c*, *serum_c* and *serum_tg*). The results in blue indicate the results for the 2014 platform and in red the ones for the platform of 2020. The label on the y-axis indicates the biobank, the total number of samples with available quantification and the Standard Deviation of the clinically measured metabolite.

### The *MetaboHealth* score shows a comparable association with mortality using re-quantified data

Next, we evaluated whether the platform changes affected the replication of the *MetaboHealth* score [2]. The *MetaboHealth* score correlated on average ∼0.83 between the 2014 platform and the re-quantification in 2020 over all the cohorts (Figure 5A); with a maximum of 0.91 (in LLS-SIBS) and a minimum of 0.72 BIOMARCS. Higher correlations for LLS-SIBS [89÷103 y.o.] and PROSPER [70÷85 y.o.] might be explained by the stronger signal caused by the fact that these cohorts generally include older individuals, with a high frequency of mortality or cardiovascular events. Cohort-specific differences in correlations between platform versions could be explained by inconsistent correlations of *acace*, *albumin*, *s_hdl_l* and *xxl_vldl_d* that have relatively high coefficients in the *MetaboHealth* score (in Figure 5B). Indeed, we notice that patient cohorts such as BIOMARCS, RAAK and UCORBIO do have lower correlations.

**Figure 5:**
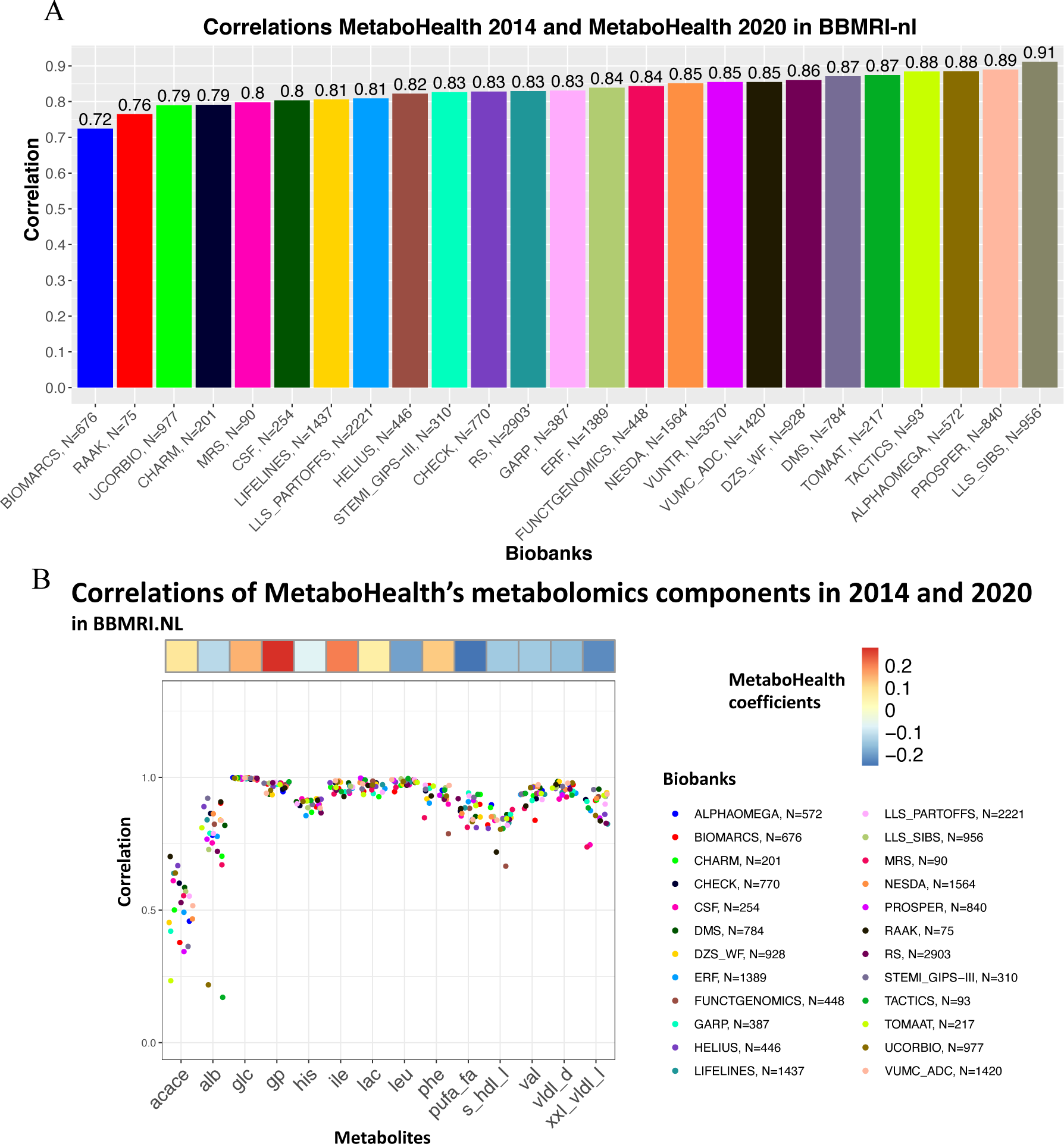
*MetaboHealth score consistency over BBMRI.NL:* [A] Bar-plot presenting the correlation of the MetaboHealth score calculated in all the BBMRI.NL biobanks with the metabolites in the data measured in 2014 or 2020; [B] Jitter-plot of the correlations of the metabolic markers used to build the MetaboHealth score calculated in data 2014 and 2020, divided per biobank. The heatmap on top shows the coefficients of each biomarker in the MetaboHealth score.

Since the *MetaboHealth* score maintained similar predictions in the platform with re-quantified metabolites, we next were interested whether the re-quantified score also showed similar associations with mortality. To this end, we modeled time-to-death using a Cox proportional Hazards model, while adjusting for age, sex and family relation, in LLS-SIBS (N_total_=797, N_events_=791). Both versions remained significantly associated (2014: HR∼2.18, p=5.42×10^−28^, and 2020: HR∼1.98, p=1×10^−30^) albeit with a slightly attenuated effect size for the 2020 platform version, but a more significant association (Figure 6).

**Figure 6:**
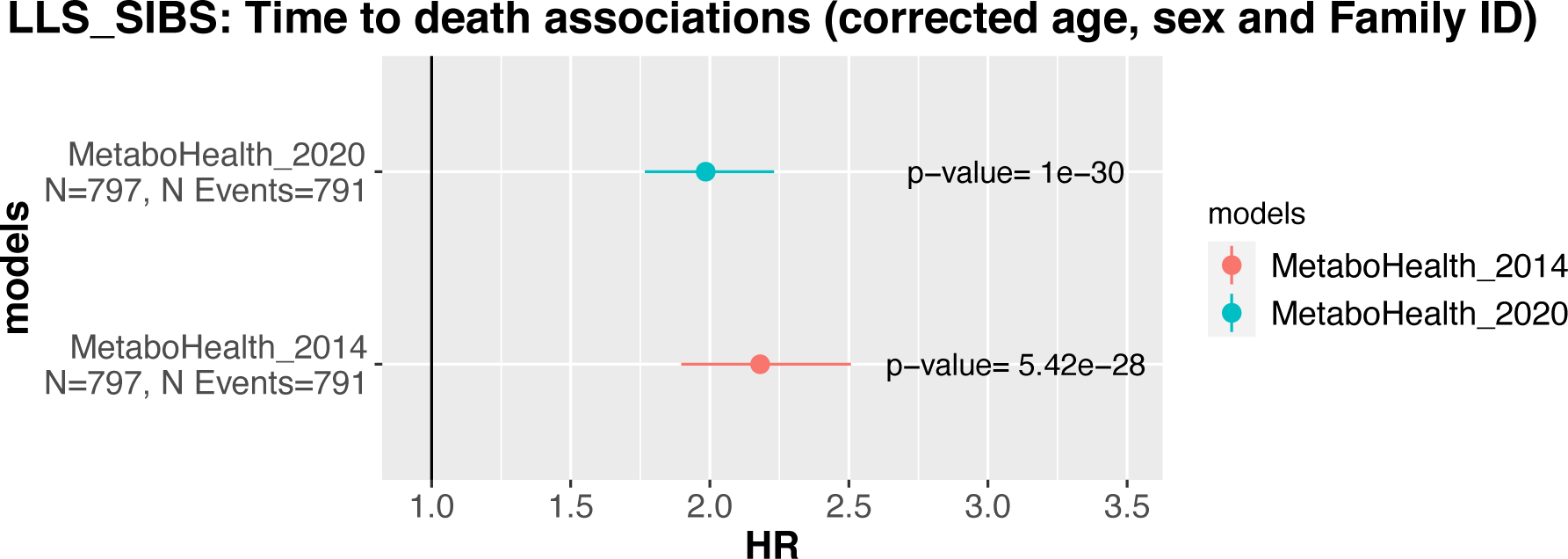
*MetaboHealth score associations with Time to Death associations in LLS-SIBS:* Association with Time to Death of the MetaboHealth score calculated with the metabolic markers quantified in 2014 (*MetaboHealth_2014*) and the metabolic markers quantified in 2020 (MetaboHealth_2020). The two Cox regression models were performed on 797 individuals with 791 reported deaths and corrected for age, sex and Family relationships.

### A retrained *MetaboAge* on re-quantified data shows similar associations with mortality compared to the previous version of *MetaboAge*

Since two essential variables (*hld2_c* and *hdl3_c*) were discontinued in the 2020 platform, the original *MetaboAge* model (`MetaboAge 1.0`) could not be computed. Therefore, we decided to retrain the *MetaboAge* model using the re-quantified Nightingale 2020 measurements, either using a: 1) a linear model (LM), consistent with the previous *MetaboAge* model; and 2) an elastic net regression (EN), regularizing the contributions of each individual metabolite. 5-Fold Cross Validation, over the BBMRI.NL dataset (∼20,366 samples, after quality control), showed overall similar accuracies, with a slight advantage for the linear model (`MetaboAge 2.0: LM`, R^2^=0.451; EN, R^2^=0.449, Figure S5). Correlations between the old and new versions of the models over all the BBMRI.NL biobanks showed cohort-specific differences, with low correlations in the RAAK cohort (R=0.5) and moderately to high correlations for the ERF and FUNCTGENOMICS cohorts (R=0.85 and 0.86, respectively) (Figure S8). Nonetheless, we observe an overall high correlation between the two novel versions of the *MetaboAge* models (R=0.99) (Figure 7B), despite that the informative metabolomics features are quite different across the three models (Figure 7A). Yet the elastic net version has a slightly higher correlation with the `MetaboAge 1.0` (`LM`: R= 0.82 and `EN`: R=0.83, Figure 7A). Nonetheless, the linear model assigns higher coefficients to only few features compared to the elastic net model (Figure 7A) (`MetaboAge 1.0` [range: −150,150], `MetaboAge 2.0`: `LM` [range: −40000, 1000000], `MetaboAge 2.0`: `EN` [range: −100, 50]).

**Figure 7:**
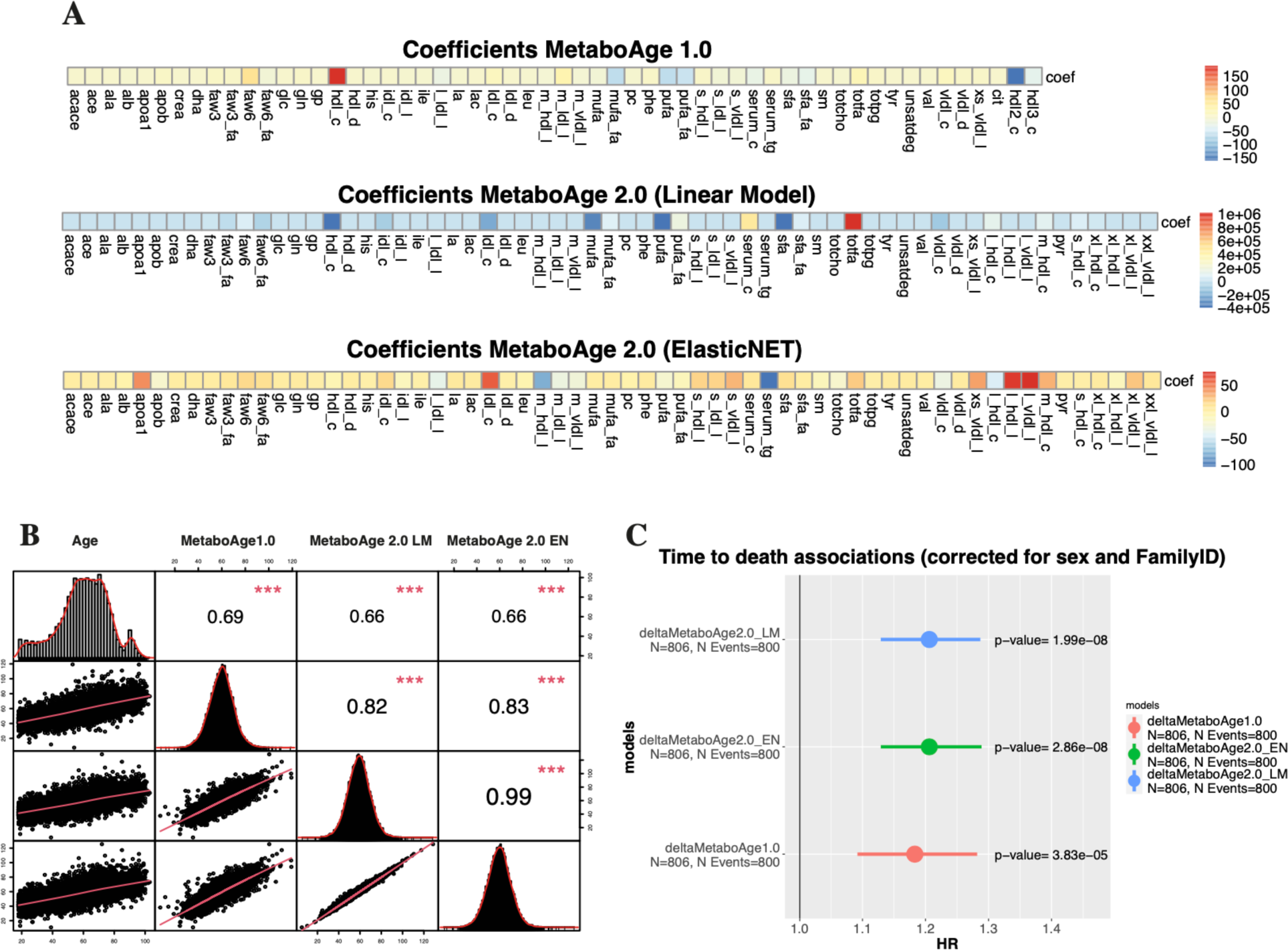
*MetaboAge 2.0 evaluations:* [A] Coefficients of MetaboAge1.0 and Metaboages2.0 ordered in the same manner; [B] Correlation between age, MetaboAge1.0, MetaboAge2.0 linear model (LM) and ElasticNET (EN); [C] Associations of time to death with the three age predictors.

Finally, we performed a Cox-regressions analysis to predict time-to-death (corrected for age sex and family relation) in the LLS-SIBS cohort (N_total_=806, N_events_=800) (Figure 7C). The associations with mortality are quite similar (equivalently significant and moderate effect sizes) across all models, but slightly higher for the `MetaboAge 2.0` models (`LM`: HR∼1.2, p=1.69×10^−08^ and `EN`: HR∼1.2, p=2.39×10^−08^, `MetaboAge 1.0:` HR∼1.18, p=2.89×10^−05^).

## Discussion

Using the BBMRI.NL biobanking consortium we evaluated the replicability across Nightingale Health platform updates between 2014, 2016, and 2020 (re-quantification). We observe improvements regarding the overall quantification quality, i.e.: a decrease in missingness; lower numbers of values that are reported as zero; and a better concordance with clinical measurements. On the other hand, there are discontinued metabolites, and changes in reported units between and sometimes within quantification versions that could affect replication efforts. Some analytes displayed low calibration correlations between the 2014/2016 and 2020 platform versions, with the 2016 version being more similar to the re-quantified data as compared to the 2014 version. Replication over the BBMRI-nl cohorts indicated similar results, however, with lower concordance for some studies (e.g., BIOMARCS, or STEMI-GIPS). Nevertheless, we identified a list of 73 variables more consistent between quantification versions across the BBMRI.NL data set (mean R>0.9). Moreover, the *MetaboHealth* score did generally replicate well between platform version in the BBMRI-nl cohorts (mean R= 0.83, min R 0.72, BIOMARCS, and max R= 0.91, LLS-SIBS). Lower correlations were attributed to inconsistencies in some score-related analytes (*acace*, *albumin*, *s_hdl_l* and *xxl_vldl_d*). Importantly, the time-to-death association of the *MetaboHealth* score was not significantly affected by the platform updates. We retrained the MetaboAge score in BBMRI-nl due to the absence of 2 analytes in the new platform version. Interestingly, the retrained version of MetaboAge recapitulated the association with time-to-death, indicating that correlations with the original MetaboAge model (`MetaboAge 1.0`) showed moderately high concordance over all cohorts in BBMRI.NL, apart for RAAK (cor∼0.5), which is a relatively small cohort of atherosclerotic people. Between the 2 versions of the MetaboAge 2.0 we believe the elastic net version to be the better model as the regularization warrants for future changes of the platform.

In conclusion, replication of previous findings and analysis of repeated measures is one of the cornerstones of epidemiological research. Hence, we call for caution when utilizing Nightingale data quantified at different time points. Moreover, is important to realize that pre-trained metabolic models cannot readily be applied across different versions of the data. In these circumstances, we recommend retraining of the score, or, if this is not possible, an extensive re-evaluation of the models and their associations with endpoints.

## Data Availability

All data produced in the present study are available upon reasonable request to BBMRI-nl.

https://www.bbmri.nl/

## Acknowledgements

This work was performed within the framework of the BBMRI Metabolomics Consortium funded by BBMRI.NL (a research infrastructure financed by the Dutch government, NWO 184.021.007 and 184.033.111), by X-omics (NWO 184.034.019), VOILA (ZonMW 457001001) and Medical Delta (scientific program METABODELTA: Metabolomics for clinical advances in the Medical Delta). EvdA is funded by a personal grant of the Dutch Research Council (NWO; VENI: 09150161810095). A full list of acknowledgements for all the contributing studies can be found in the Supplementary Materials (BBMRI.NL Cohort description).

## Contributors

EbvDA, DB, MJTR, PES and MB conceived and wrote the manuscript. DB performed the analyses. DB and MB verified the underlying data. EBvDA and MJTR verified and supervised the analyses. All authors discussed the results and contributed to the final manuscript. All authors read and approved the final version of the manuscript.

## Data and code availability

The data is available upon request at https://www.bbmri.nl/. A presentation of the results with the code to reproduce this work can be found at (https://github.com/DanieleBizzarri/NightingaleMetabolomics_Requantification2020).

## Materials and Methods

### 1. Dataset descriptions

The Dutch Biobanking and BioMolecular resources and Research Infrastructure (BBMRI.NL) is a large consortium composed of 28 Dutch cohorts, which quantified their samples with the Nightingale Health platform in different time points, allowing an investigation on the platform differences over the years. About 25,000 samples from 26 cohorts were quantified during the first wave in 2014. A second wave of 10,000 samples was then obtained in 2016, including some longitudinal time-points and 2 new cohorts. Finally, after the 2020 update of the platform, the entire BBMRI.NL (35,000 samples from 28 cohorts) was re-quantified to have comparable measures to other Consortia.

#### A. BBMRI.NL

BBMRI.NL (https://www.bbmri.nl/) is a Dutch Consortium which includes a total of 35,000 samples from the following 28 Dutch biobanks: ALPHAOMEGA, BIOMARCS, CHARM, CHECK, CODAM, CSF, DMS, DZS_WF, ERF, FUNCTGENOMICS, GARP, HELIUS, HOF, LIFELINES, LLS_PARTOFFS, LLS_SIBS, MRS, NESDA, PROSPER, RAAK, RS, STABILITEIT, STEMI_GIPS-III, TACTICS, TOMAAT, UCORBIO, VUMC_ADC, VUNTR. Complete descriptions and ethics statement of each cohort is added to the Supplementary Material.

**Metabolomics Dataset:** Nightingale Health performed the quantification of high throughput proton Nuclear Magnetic Resonance (^1^H-NMR) for the EDTA plasma for BBMRI.NL in separate waves (***Table 1***). The first wave was performed in 2014, on a great portion of the data (∼25,000 samples). The second wave was performed in 2016 to quantify ^1^H-NMR metabolomics in the cohorts HOF and STABILITEIT, but also to quantify follow-ups sampling from different cohorts. Finally, in 2021 a re-quantification was performed almost to the entire dataset to update the metabolomics measurements to the latest platform version (platform version 2020).

#### B. The Leiden Longevity Study

The Leiden Longevity Study is one of the cohort included in BBMRI.NL, which comprises a first generation subgroup of long-lived parents (LLS-SIBS, age= 89 ÷ 103 years old) and a second generation which includes their middle-aged offspring with the relative partner (LLS-PAROFFS age median = 30 ÷ 79 years old) [11].

**Metabolomics Dataset:** While only one sample collection was performed on the older individuals of LLS-SIBS [998 individuals], there are three time-points available for LLS-PAROFFS drawn with ∼3 years gap one after the other (IOP1, IOP2 and IOP3) (Table 1). The first-time point (IOP1, 2,313 individuals) was quantified during the first wave in 2014, while the second and third samples measurements (IOP2 and IOP3, respectively 670 and 498 individuals) were included in the second wave, with the platform version 2016. All the samples were then re-quantified in 2021 with the rest of BBMRI.NL data. The last column of the table shows the number of common samples after the quality control of the two datasets, described in the next paragraph.

### 2. Comparison of the metabolomic analytes

**Preprocessing:** All the three versions of the metabolomics assays were run by Nightingale Health on EDTA-plasma samples handled by the BBMRI.NL cohorts. More than 220 analytes are included in all nightingale platform, however we decided to mostly focus our attention on the 63 mutually independent analytes used to build the previous metabolomics-based models [2,6,12]. However, since 2 of these analytes were discontinued (hdl2_c and hdl3_c), we substituted them with 4 biologically equivalent analytes, upon Nightingale’s Health advice (xl_hdl_c, l_hdl_c, m_hdl_c, s_hdl_c) (lists in Supplementary Materials), which are available in all datasets. We then removed samples with more than 1 missing value, more than one zero and more than one outlier, defined as having a concentration more than 5 standard deviations away from the mean of the analyte.

**Analyses:** We used Spearman’s correlation to measure the strength and direction of monotonic associations between the analytes in the different versions of the platform. We also used a Median Absolute Distance to evaluate the error of Nightingale Health’s analytes to the clinically measured values. The Mean absolute distance is obtained by using mean and standard deviations of the clinical measures to scale all measures (both clinical and Nightingale quantifications) to have comparable results.

### 3. MetaboHealth Score

**Preprocessing:** The MetaboHealth score was applied to both the datasets (the first wave and the re-quantified), according to the description by Deelen et al. [2], using the R-package MiMIR [13]. First, a logarithm transformation was applied to the analytes, while adding a value of 1 to all analytes containing any zero. A z-scale normalization was then applied to the log-transformed analytes in each cohort separately. Finally, the coefficients as indicated by Deelen et al. [2] were applied to the dataset.

**Analyses:** Once we obtained the score, we used spearman’s correlation to compare the differences in MetaboHealth score before and after re-quantification. Cox proportional hazard models are then used to test the associations between the two MetaboHealth scores and time to death.

### 4. MetaboAge

**Preprocessing:** The quality control process used for the dataset in the first wave of measures (data 2014) is discussed in details in our previous publications [6,12]. We used the same steps also in the re-quantified dataset. From the above-mentioned list of 65 analytes, we decided not to consider analytes with low detection rates in several cohorts (citrate and 3-hydroxybutyrate). We then excluded cohorts with several problems in the 65 selected analytes. VUNTR (N=3559) has high levels of missingness in pyruvate and glutamine, while CODAM (N=145) presented outliers in several metabolic features. We also removed samples with 1 or more missing value (65 samples), one or more zeroes per sample (1 sample) and one or more concentration more than 5 times the standard deviations away from the general mean of the feature (644 samples). The remaining 265 missing values (0.021% of the remaining values) were imputed using nipals (in the R package pcaMethods). The final dataset, comprising 20,366 samples and 63 analytes, was z-scaled to have comparable concentrations across all features.

**Analyses:** Due to discontinued analytes, we had to retrain the models and we decided to train 2 different types of models: a linear regression model, to maintain the model as close as possible to the previous version, and an ElasticNET regression, which avoids overfitting thanks to a regularization technique. To train and evaluate both models we employed a 5-Fold Cross Validation scheme. During the training of the ElasticNET model we fixed the mixing parameter α to 0.5 and optimized the shrinkage parameter λ (like it was done in previous papers [6,14,15]). As for the MetaboHealth, we then used spearman’s correlations to compare the different models and Cox proportional hazard models to investigate the associations with time to death.

## References

[1] van den Akker Erik B., Trompet Stella, Barkey Wolf Jurriaan J.H., Beekman Marian, Suchiman H. Eka D., Deelen Joris, et al. Metabolic Age Based on the BBMRI-NL 1H-NMR Metabolomics Repository as Biomarker of Age-related Disease. Circulation: Genomic and Precision Medicine 2020;13:541–7. https://doi.org/10.1161/CIRCGEN.119.002610.

[2] Deelen J, Kettunen J, Fischer K, van der Spek A, Trompet S, Kastenmüller G, et al. A metabolic profile of all-cause mortality risk identified in an observational study of 44,168 individuals. Nature Communications 2019;10:1–8. https://doi.org/10.1038/s41467-019-11311-9.

[3] Nightingale Health UK Biobank Initiative, Julkunen H, Cichońska A, Slagboom PE, Würtz P. Metabolic biomarker profiling for identification of susceptibility to severe pneumonia and COVID-19 in the general population. ELife 2021;10:e63033. https://doi.org/10.7554/eLife.63033.

[4] Buergel T, Steinfeldt J, Ruyoga G, Pietzner M, Bizzarri D, Vojinovic D, et al. Metabolomic profiles predict individual multidisease outcomes. Nat Med 2022;28:2309–20. https://doi.org/10.1038/s41591-022-01980-3.

[5] Group NHBC, Barrett JC, Esko T, Fischer K, Jostins-Dean L, Jousilahti P, et al. Metabolomic and genomic prediction of common diseases in 477,706 participants in three national biobanks 2023:2023.06.09.23291213. https://doi.org/10.1101/2023.06.09.23291213.

[6] Bizzarri D, Reinders MJT, Beekman M, Slagboom PE, Bbmri-nl, van den Akker EB. 1H-NMR metabolomics-based surrogates to impute common clinical risk factors and endpoints. EBioMedicine 2022;75:103764. https://doi.org/10.1016/j.ebiom.2021.103764.

[7] Bharti SK, Roy R. Quantitative 1H NMR spectroscopy. TrAC Trends in Analytical Chemistry 2012;35:5–26. https://doi.org/10.1016/j.trac.2012.02.007.

[8] Yu B, Zanetti KA, Temprosa M, Albanes D, Appel N, Barrera CB, et al. The Consortium of Metabolomics Studies (COMETS): Metabolomics in 47 Prospective Cohort Studies. Am J Epidemiol 2019;188:991–1012. https://doi.org/10.1093/aje/kwz028.

[9] Singh T, Kurki MI, Curtis D, Purcell SM, Crooks L, McRae J, et al. Rare loss-of-function variants in SETD1A are associated with schizophrenia and developmental disorders. Nat Neurosci 2016;19:571–7. https://doi.org/10.1038/nn.4267.

[10] Julkunen H, Cichońska A, Tiainen M, Koskela H, Nybo K, Mäkelä V, et al. Atlas of plasma NMR biomarkers for health and disease in 118,461 individuals from the UK Biobank. Nat Commun 2023;14:604. https://doi.org/10.1038/s41467-023-36231-7.

[11] Schoenmaker M, de Craen AJM, de Meijer PHEM, Beekman M, Blauw GJ, Slagboom PE, et al. Evidence of genetic enrichment for exceptional survival using a family approach: the Leiden Longevity Study. European Journal of Human Genetics 2006;14:79–84. https://doi.org/10.1038/sj.ejhg.5201508.

[12] van den Akker Erik B., Trompet Stella, Barkey Wolf Jurriaan J.H., Beekman Marian, Suchiman H. Eka D., Deelen Joris, et al. Metabolic Age Based on the BBMRI-NL 1H-NMR Metabolomics Repository as Biomarker of Age-related Disease. Circulation: Genomic and Precision Medicine n.d.;0. https://doi.org/10.1161/CIRCGEN.119.002610.

[13] Bizzarri D, Reinders MJT, Beekman M, Slagboom PE, van den Akker EB. MiMIR: R-shiny application to infer risk factors and endpoints from Nightingale Health’s 1H-NMR metabolomics data. Bioinformatics 2022;38:3847–9. https://doi.org/10.1093/bioinformatics/btac388.

[14] Horvath S. DNA methylation age of human tissues and cell types. Genome Biol 2013;14:R115. https://doi.org/10.1186/gb-2013-14-10-r115.

[15] Lu AT, Quach A, Wilson JG, Reiner AP, Aviv A, Raj K, et al. DNA methylation GrimAge strongly predicts lifespan and healthspan. Aging (Albany NY) 2019;11:303–27. https://doi.org/10.18632/aging.101684.

