## Supplementary Materials for "Technical report: A comprehensive comparison between different quantification versions of Nightingale Health’s ^1^H-NMR metabolomics platform"

### Supplementary Figures

#### Figure S1

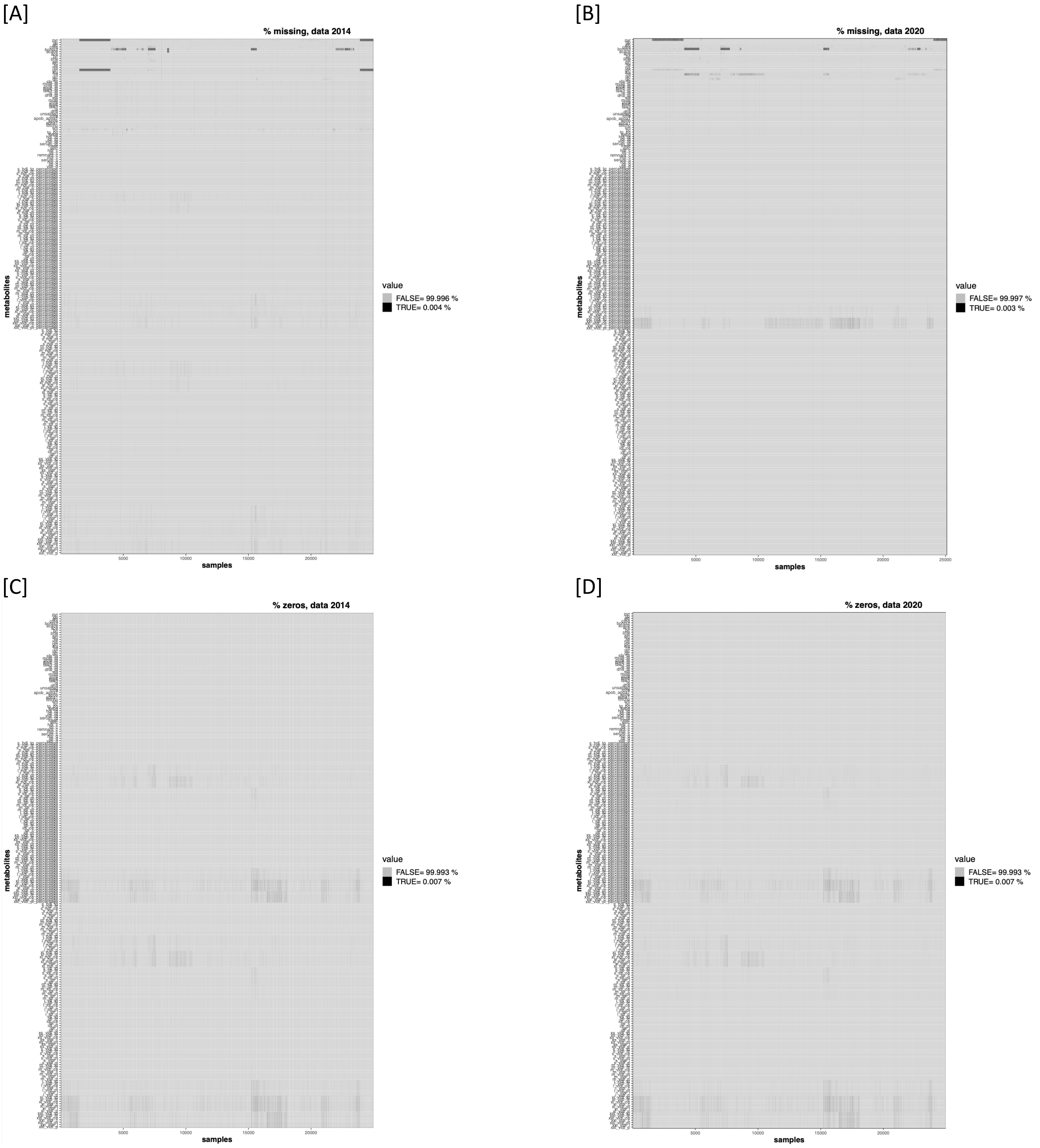

[E]

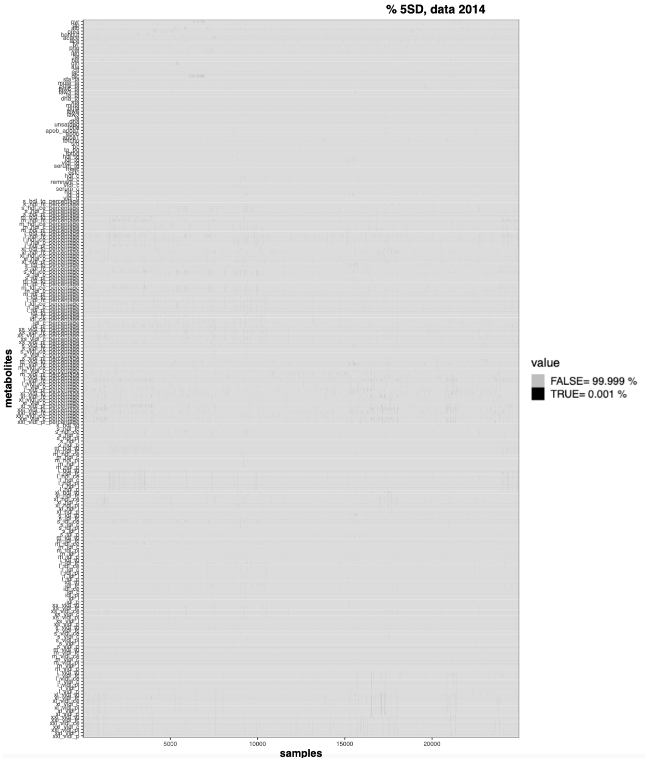

[F]

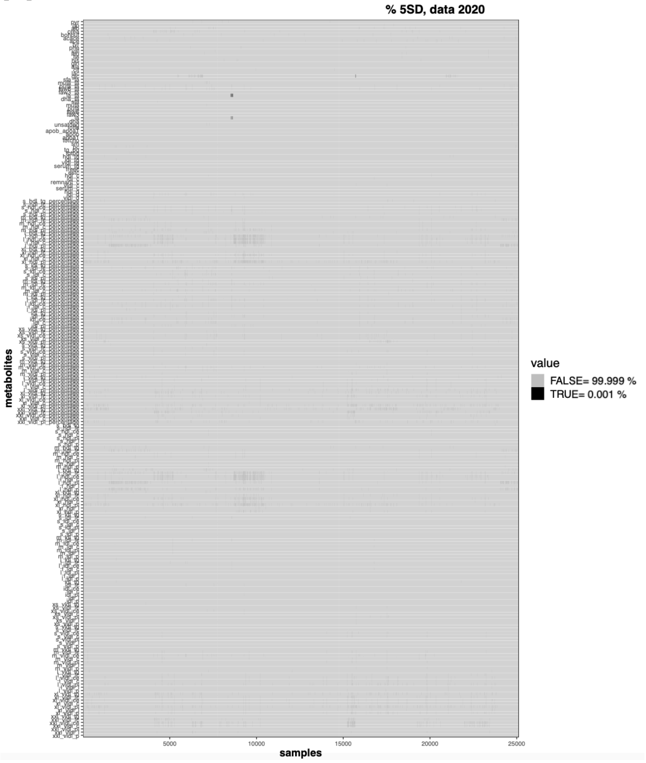

**Figure S1:** Heatmaps reporting in black the samples with [A-B] missing, [C-D] zeros, and [E-F] outliers in the Nightingale Health metabolomics dataset of 2014 and 2020.

#### Figure S2

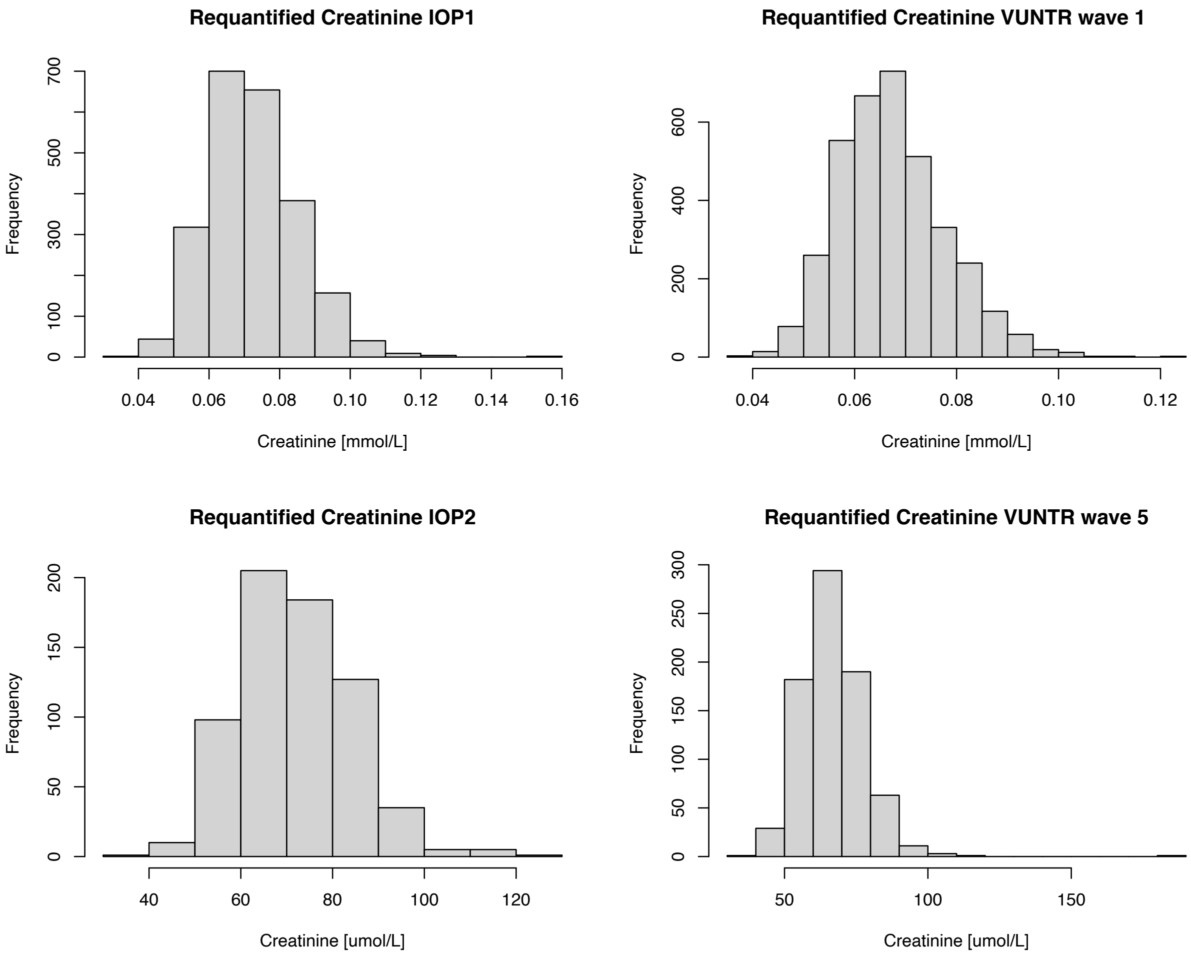

**Figure S2:** Distribution of creatinine in the Nightingale Metabolomics dataset of 2020. On the left the distributions in LLS-PAROFFS IOP1 (top), which was quantified the first time in 2014 and the repeated measures of IOP2 (bottom), quantified the first time in 2016. On the right wave 1 (top), quantified the first time in 2014, and wave 5 (bottom), quantified the first time in 2016, of VUNTR.

Figure S3

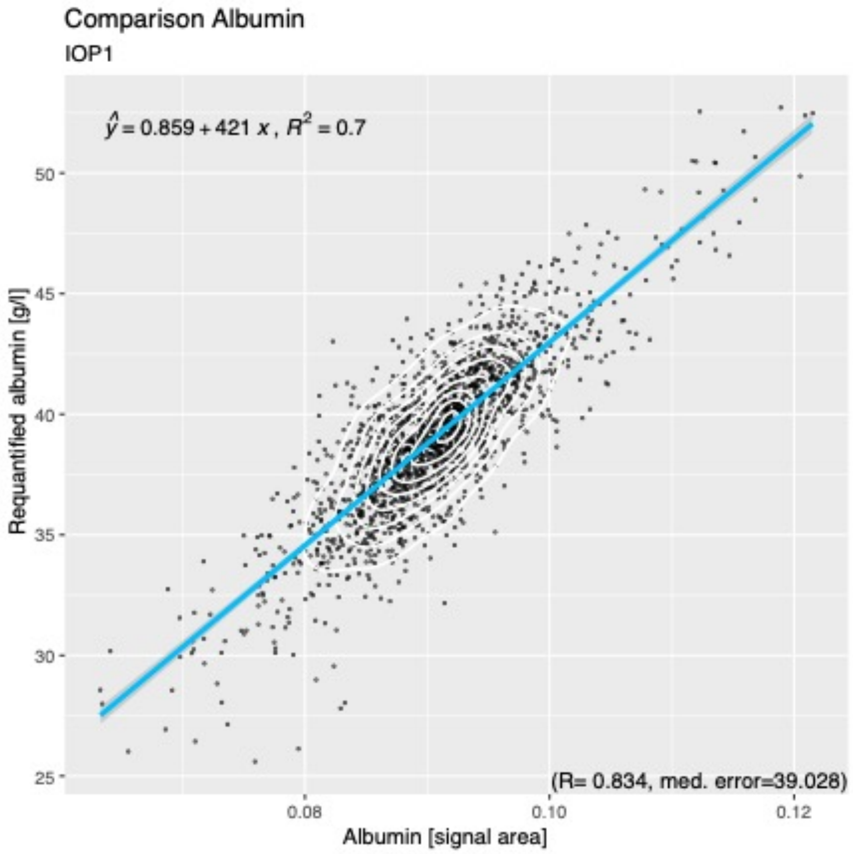

**Figure S3:** Comparison Albumin measured in the first wave (x-axis) and re-quantified (y-axis), with axes adjusted to the ranges of each variable.

#### Figure S4

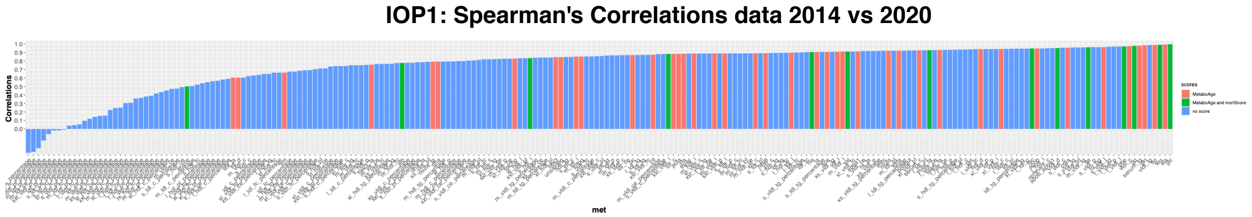

**Figure S4:** Spearman's correlations comparing the 2014 and 2020 versions of all 220 metabolomics features.

#### Figure S5

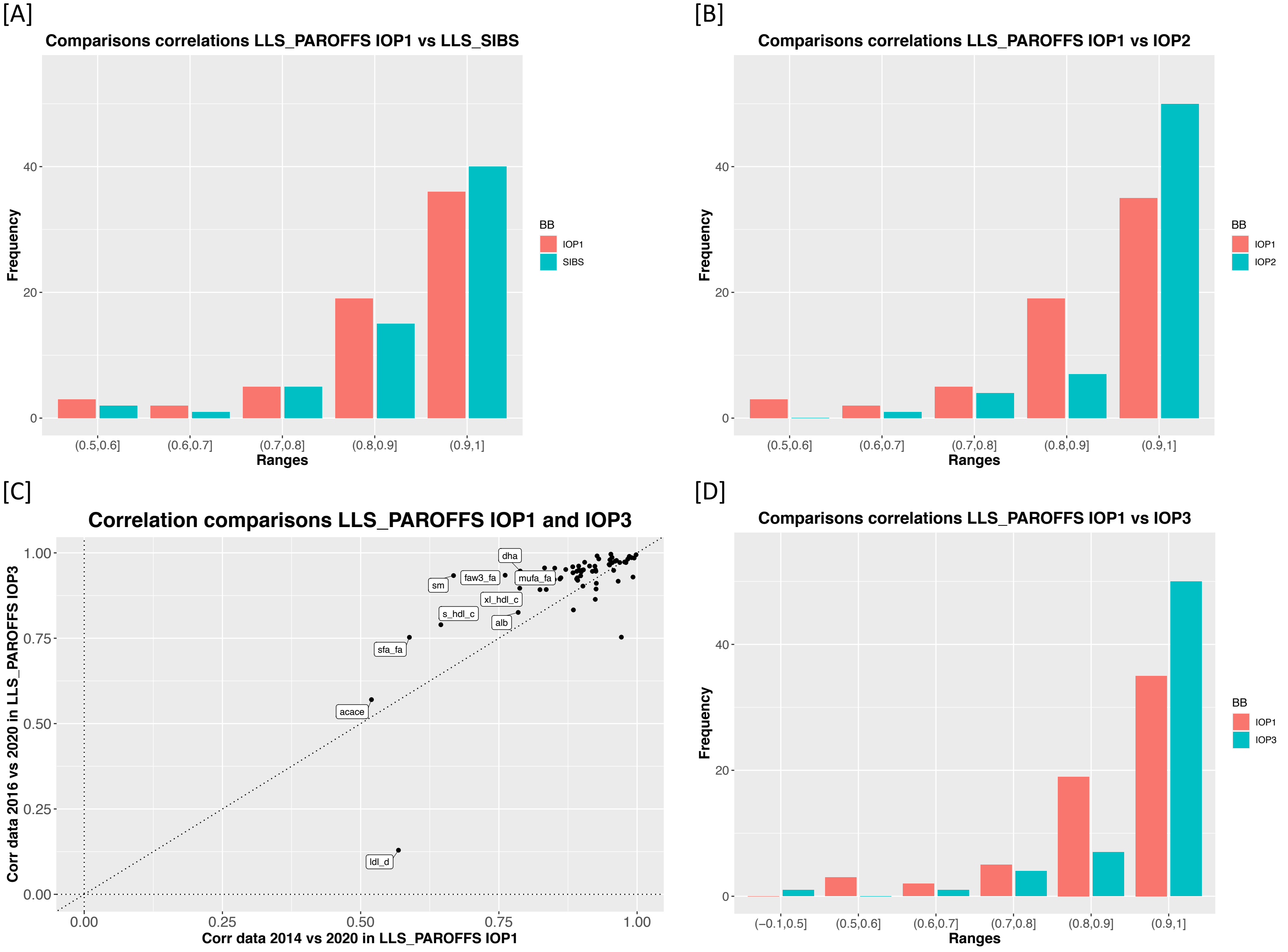

***Figure S5:*** *Comparisons of the correlations of the metabolites before and after re-quantification in LLS-SIBS and LLS-PAROFFS IOP1,2 and 3:* [A] Bar-plot comparing of the correlations of the metabolites in IOP1 and LLS-SIBS divided in ranges,

[B] Bar-plot comparing of the correlations in LLS-PAROFFS IOP1 and 2, [C] Scatterplot comparing the correlations of LLS-PAROFFS IOP1 and IOP3 and [C] Boxplot of the correlations in LLS-PAROFFS IOP1 and IOP3 divided in ranges.

#### Figure S6

[A]

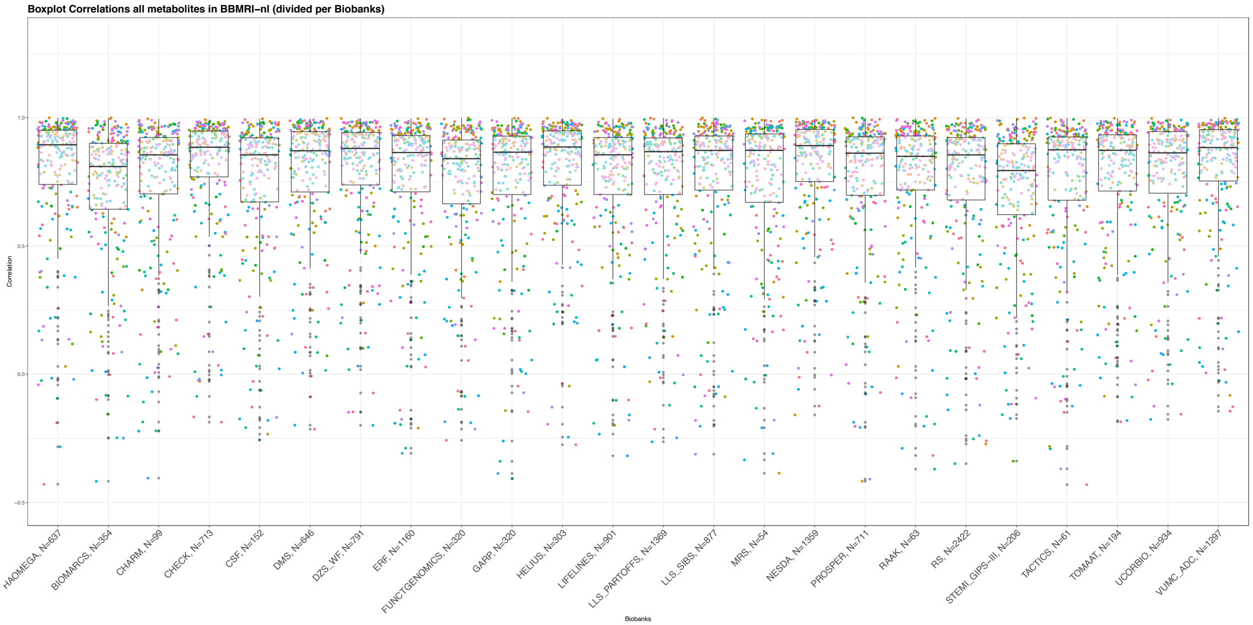

[B]

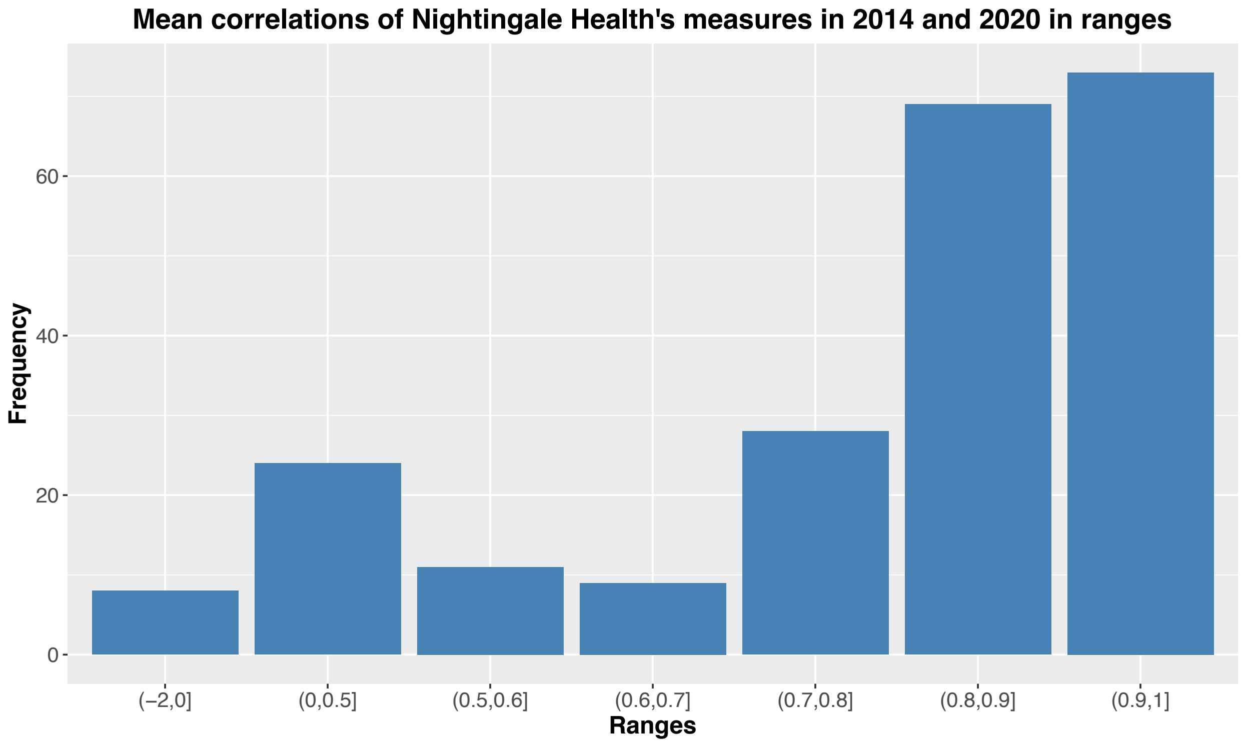

[C]

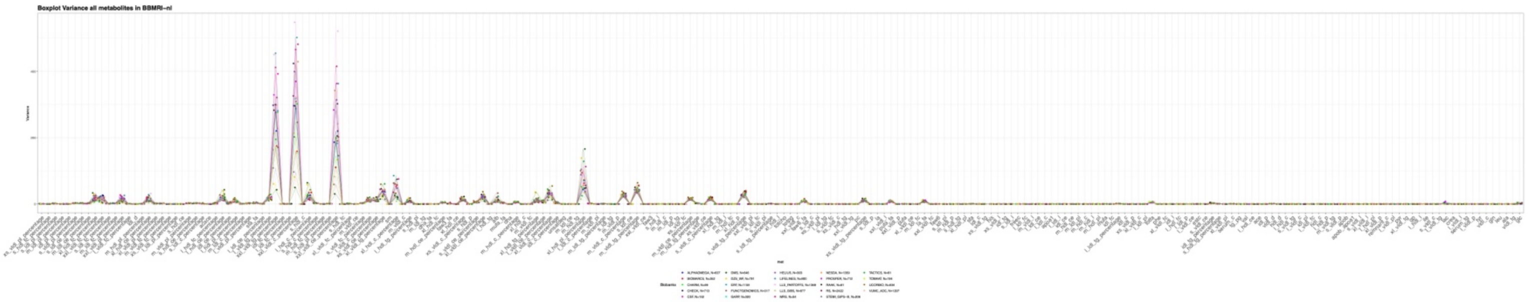

[D]

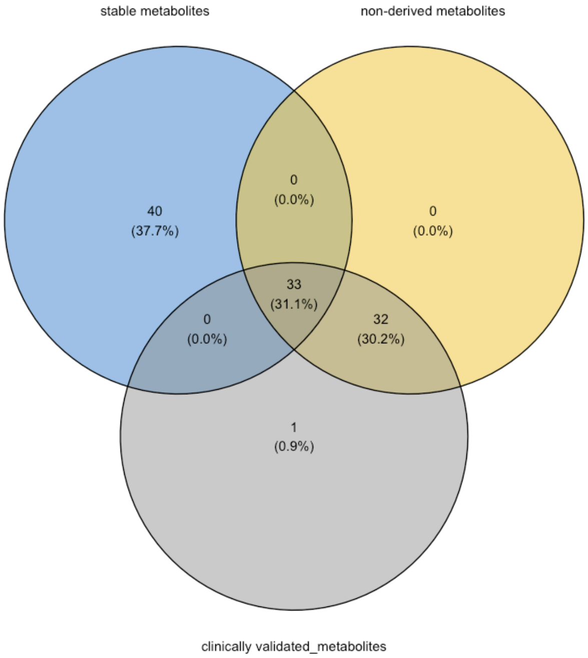

**Figure S6:** Correlations of all 220 features in all the biobanks of BBMRI-nl before and after the re-quantifications divided [A] in the biobanks and [B] divided in ranges, [C] variance of the 2020 versions of all the features in all the biobanks in BBMRI-nl, [D] Venn diagram containing the most stable metabolites over the different platforms, the 65 non-derived measures that we defined to obtain our metabolomics-based scores, and the clinically validated features.

### Figure S7

[A]

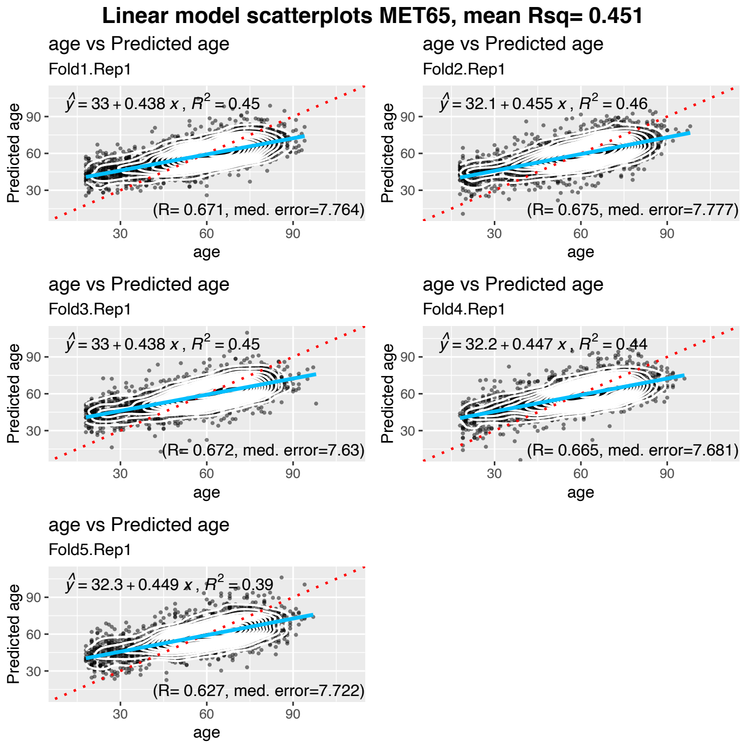
Linear model scatterplots MET65, mean Rsq= 0.451
age vs Predicted age
Fold1.Rep1
$\hat{y} = 33 + 0.438\ x,\ R^2 = 0.45$
(R= 0.671, med. error=7.764)
age vs Predicted age
Fold2.Rep1
$\hat{y} = 32.1 + 0.455\ x,\ R^2 = 0.46$
(R= 0.675, med. error=7.777)
age vs Predicted age
Fold3.Rep1
$\hat{y} = 33 + 0.438\ x,\ R^2 = 0.45$
(R= 0.672, med. error=7.63)
age vs Predicted age
Fold4.Rep1
$\hat{y} = 32.2 + 0.447\ x,\ R^2 = 0.44$
(R= 0.665, med. error=7.681)
age vs Predicted age
Fold5.Rep1
$\hat{y} = 32.3 + 0.449\ x,\ R^2 = 0.39$
(R= 0.627, med. error=7.722)
Predicted age
age
30
60
90

[B]

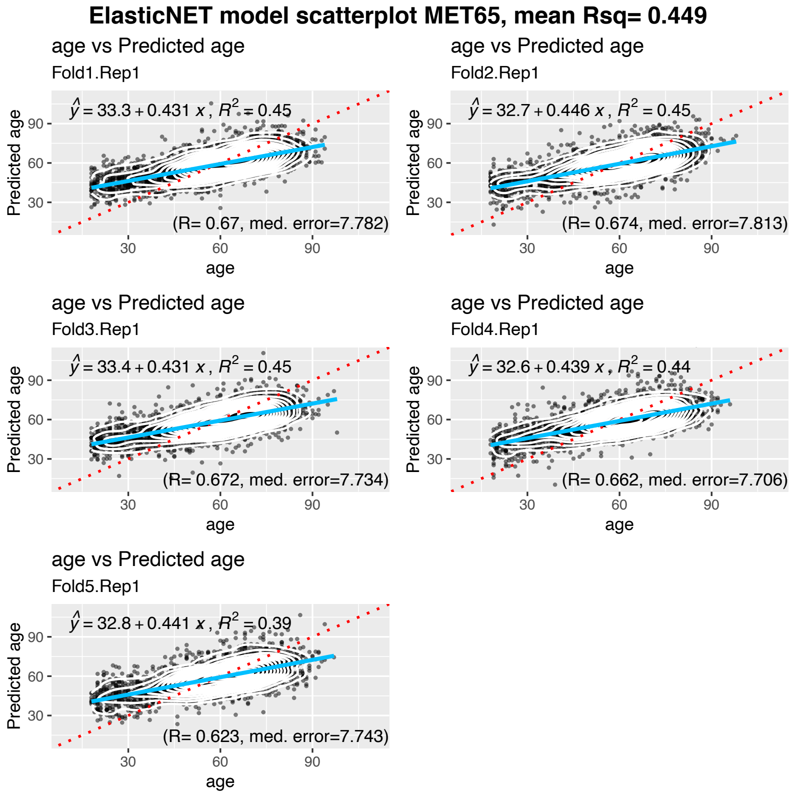
ElasticNET model scatterplot MET65, mean Rsq= 0.449
age vs Predicted age
Fold1.Rep1
$\hat{y} = 33.3 + 0.431\ x,\ R^2 = 0.45$
(R= 0.67, med. error=7.782)
age vs Predicted age
Fold2.Rep1
$\hat{y} = 32.7 + 0.446\ x,\ R^2 = 0.45$
(R= 0.674, med. error=7.813)
age vs Predicted age
Fold3.Rep1
$\hat{y} = 33.4 + 0.431\ x,\ R^2 = 0.45$
(R= 0.672, med. error=7.734)
age vs Predicted age
Fold4.Rep1
$\hat{y} = 32.6 + 0.439\ x,\ R^2 = 0.44$
(R= 0.662, med. error=7.706)
age vs Predicted age
Fold5.Rep1
$\hat{y} = 32.8 + 0.441\ x,\ R^2 = 0.39$
(R= 0.623, med. error=7.743)
Predicted age
age
30
60
90

**Figure S7:** 5-Fold Cross-Validation results for the MetaboAge 2.0 for [A] the linear model and [B] the ElasticNET model.

#### Figure S8

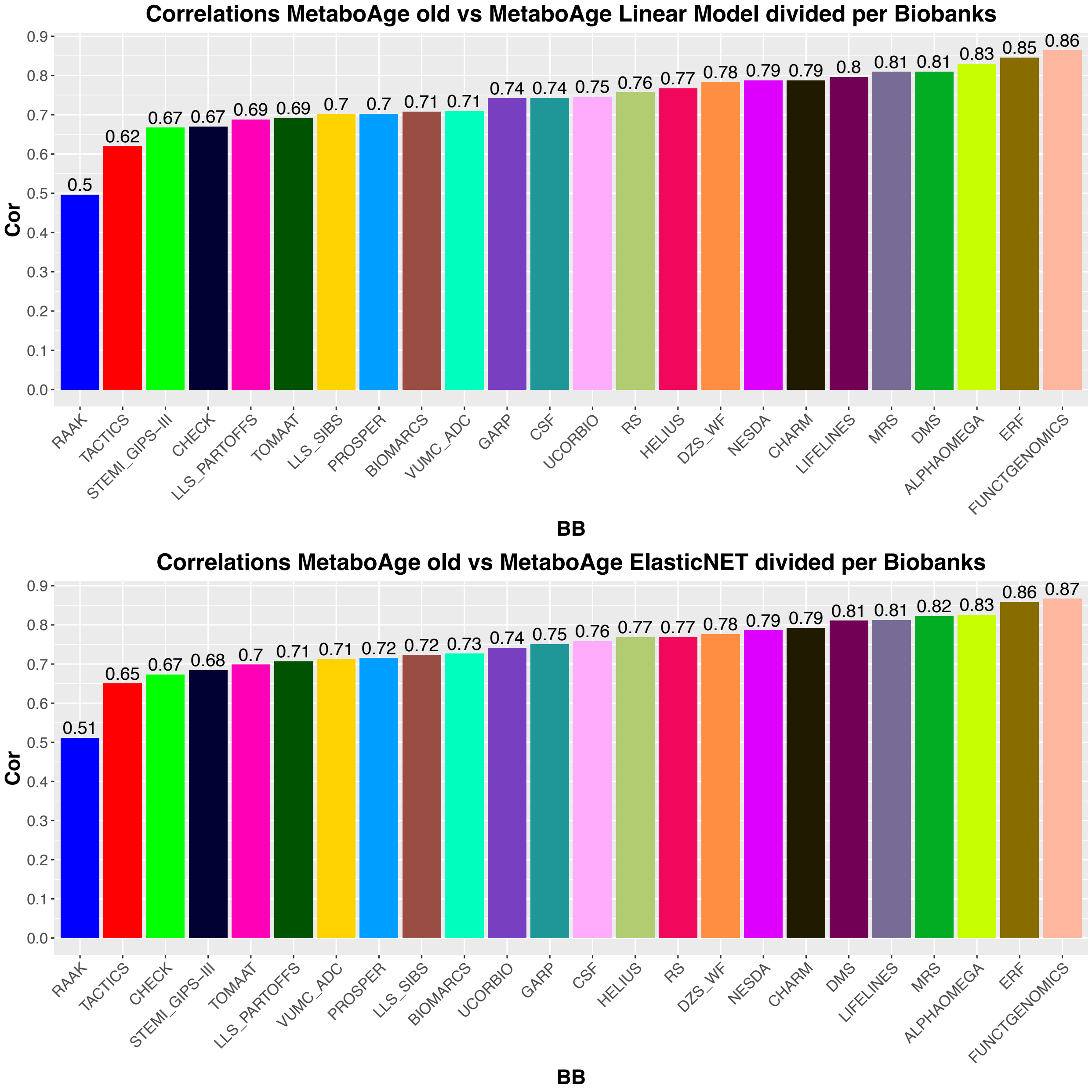

**Figure S8:** Correlations of MetaboAge 1.0 and MetaboAge 2.0 ([A] linear model and [B] ElasticNET) across all the cohorts in BBMRI-nl.

### Metabolites lists

#### 1. List of the 65 metabolites used to build the novel MetaboAge

| name | BBMRI_names | description |
|---|---|---|
| Total-C | serum_c | Total cholesterol |
| VLDL-C | vldl_c | VLDL cholesterol |
| LDL-C | ldl_c | LDL cholesterol |
| HDL-C | hdl_c | HDL cholesterol |
| Total triglycerides | serum_tg | Total triglycerides |
| VLDL particle size | vldl_d | Average diameter for VLDL particles |
| LDL particle size | ldl_d | Average diameter for LDL particles |
| HDL particle size | hdl_d | Average diameter for HDL particles |
| Phosphoglycerides | totpg | Phosphoglycerides |
| Total cholines | totcho | Total cholines |
| Phosphatidylcholines | pc | Phosphatidylcholines |
| Sphingomyelins | sm | Sphingomyelins |
| ApoB | apob | Apolipoprotein B |
| ApoA1 | apoa1 | Apolipoprotein A1 |
| Total fatty acids | totfa | Total fatty acids |
| Unsaturation | unsatdeg | Degree of unsaturation |
| Omega-3 | faw3 | Omega-3 fatty acids |
| Omega-6 | faw6 | Omega-6 fatty acids |
| PUFA | pufa | Polyunsaturated fatty acids |
| MUFA | mufa | Monounsaturated fatty acids |
| SFA | sfa | Saturated fatty acids |
| LA | la | Linoleic acid |
| DHA | dha | Docosahexaenoic acid |
| Omega-3 % | faw3_fa | Ratio of omega-3 fatty acids to total fatty acids |
| Omega-6 % | faw6_fa | Ratio of omega-6 fatty acids to total fatty acids |
| PUFA % | pufa_fa | Ratio of polyunsaturated fatty acids to total fatty acids |
| MUFA % | mufa_fa | Ratio of monounsaturated fatty acids to total fatty acids |
| SFA % | sfa_fa | Ratio of saturated fatty acids to total fatty acids |
| Alanine | ala | Alanine |
| Glutamine | gln | Glutamine |
| Histidine | his | Histidine |
| Isoleucine | ile | Isoleucine |
| Leucine | leu | Leucine |
| Valine | val | Valine |
| Phenylalanine | phe | Phenylalanine |
| Tyrosine | tyr | Tyrosine |
| Glucose | glc | Glucose |

| | | |
|---|---|---|
| Lactate | lac | Lactate |
| Pyruvate | pyr | Pyruvate |
| Citrate | cit | Citrate |
| 3-Hydroxybutyrate | bohbut | 3-Hydroxybutyrate |
| Acetate | ace | Acetate |
| Acetoacetate | acace | Acetoacetate |
| Creatinine | crea | Creatinine |
| Albumin | alb | Albumin |
| Glycoprotein acetyls | gp | Glycoprotein acetyls |
| XXL-VLDL-L | xxl_vldl_l | Total lipids in chylomicrons and extremely large VLDL |
| XL-VLDL-L | xl_vldl_l | Total lipids in very large VLDL |
| L-VLDL-L | l_vldl_l | Total lipids in large VLDL |
| M-VLDL-L | m_vldl_l | Total lipids in medium VLDL |
| S-VLDL-L | s_vldl_l | Total lipids in small VLDL |
| XS-VLDL-L | xs_vldl_l | Total lipids in very small VLDL |
| IDL-L | idl_l | Total lipids in IDL |
| IDL-C | idl_c | Cholesterol in IDL |
| L-LDL-L | l_ldl_l | Total lipids in large LDL |
| M-LDL-L | m_ldl_l | Total lipids in medium LDL |
| S-LDL-L | s_ldl_l | Total lipids in small LDL |
| XL-HDL-L | xl_hdl_l | Total lipids in very large HDL |
| XL-HDL-C | xl_hdl_c | Cholesterol in very large HDL |
| L-HDL-L | l_hdl_l | Total lipids in large HDL |
| L-HDL-C | l_hdl_c | Cholesterol in large HDL |
| M-HDL-L | m_hdl_l | Total lipids in medium HDL |
| M-HDL-C | m_hdl_c | Cholesterol in medium HDL |
| S-HDL-L | s_hdl_l | Total lipids in small HDL |
| S-HDL-C | s_hdl_c | Cholesterol in small HDL |

#### 2. Discontinued metabolites in the platform 2020

| name | BBMRI_names | description |
|---|---|---|
| HDL2-C | hdl2_c | HDL2 cholesterol |
| HDL3-C | hdl3_c | HDL3 cholesterol |

#### 3. Metabolites added to the Nightingale platform with the update of 2020

| Name | BBMRI_names | description |
|---|---|---|
| non-HDL-C | non_hdl_c | Total cholesterol minus HDL-C |
| Clinical LDL-C | clinical_ldl_c | Clinical LDL cholesterol |
| Total-PL | total_pl | Total phospholipids in lipoprotein particles |
| VLDL-PL | vldl_pl | Phospholipids in VLDL |

| | | |
|---|---|---|
| LDL-PL | ldl_pl | Phospholipids in LDL |
| HDL-PL | hdl_pl | Phospholipids in HDL |
| VLDL-CE | vldl_ce | Cholesteryl esters in VLDL |
| LDL-CE | ldl_ce | Cholesteryl esters in LDL |
| VLDL-FC | vldl_fc | Free cholesterol in VLDL |
| LDL-FC | ldl_fc | Free cholesterol in LDL |
| HDL-FC | hdl_fc | Free cholesterol in HDL |
| Total-L | total_l | Total lipids in lipoprotein particles |
| VLDL-L | vldl_l | Total lipids in VLDL |
| LDL-L | ldl_l | Total lipids in LDL |
| HDL-L | hdl_l | Total lipids in HDL |
| Total-P | total_p | Total concentration of lipoprotein particles |
| VLDL-P | vldl_p | Concentration of VLDL particles |
| LDL-P | ldl_p | Concentration of LDL particles |
| HDL-P | hdl_p | Concentration of HDL particles |
| PUFA/MUFA | pufa_by_mufa | Ratio of polyunsaturated fatty acids to monounsaturated fatty acids |
| Omega-6/Omega-3 | faw6_by_faw3 | Ratio of omega-6 fatty acids to omega-3 fatty acids |
| Total BCAA | total_bcaa | Total concentration of branched-chain amino acids (leucine + isoleucine + valine) |
| Acetone | acetone | Acetone |

#### 4. Clinically validated metabolites

| name | BBMRI_names | description |
|---|---|---|
| Total-C | serum_c | Total cholesterol |
| VLDL-C | vldl_c | VLDL cholesterol |
| Clinical LDL-C | clinical_ldl_c | Clinical LDL cholesterol |
| HDL-C | hdl_c | HDL cholesterol |
| Total triglycerides | serum_tg | Total triglycerides |
| ApoB | apob | Apolipoprotein B |
| ApoA1 | apoa1 | Apolipoprotein A1 |
| ApoB/ApoA1 | apob_apoa1 | Ratio of apolipoprotein B to apolipoprotein A1 |
| Total fatty acids | totfa | Total fatty acids |
| Omega-3 | faw3 | Omega-3 fatty acids |
| Omega-6 | faw6 | Omega-6 fatty acids |
| PUFA | pufa | Polyunsaturated fatty acids |
| MUFA | mufa | Monounsaturated fatty acids |
| SFA | sfa | Saturated fatty acids |
| DHA | dha | Docosahexaenoic acid |
| Omega-3 % | faw3_fa | Ratio of omega-3 fatty acids to total fatty acids |
| Omega-6 % | faw6_fa | Ratio of omega-6 fatty acids to total fatty acids |

| | | |
|---|---|---|
| PUFA % | pufa_fa | Ratio of polyunsaturated fatty acids to total fatty acids |
| MUFA % | mufa_fa | Ratio of monounsaturated fatty acids to total fatty acids |
| SFA % | sfa_fa | Ratio of saturated fatty acids to total fatty acids |
| DHA % | dha_fa | Ratio of docosahexaenoic acid to total fatty acids |
| PUFA/MUFA | pufa_by_mufa | Ratio of polyunsaturated fatty acids to monounsaturated fatty acids |
| Omega-6/Omega-3 | faw6_by_faw3 | Ratio of omega-6 fatty acids to omega-3 fatty acids |
| Alanine | ala | Alanine |
| Glycine | glycine | Glycine |
| Histidine | his | Histidine |
| Total BCAA | total_bcaa | Total concentration of branched-chain amino acids (leucine + isoleucine + valine) |
| Isoleucine | ile | Isoleucine |
| Leucine | leu | Leucine |
| Valine | val | Valine |
| Phenylalanine | phe | Phenylalanine |
| Tyrosine | tyr | Tyrosine |
| Glucose | glc | Glucose |
| Lactate | lac | Lactate |
| Creatinine | crea | Creatinine |
| Albumin | alb | Albumin |

#### 5. 73 stable metabolites

| name | BBMRI_names | description |
|---|---|---|
| L-LDL-CE | l_ldl_ce | Total cholesterol |
| L-LDL-C | l_ldl_c | Remnant cholesterol (non-HDL, non-LDL -cholesterol) |
| ApoA1 | apoa1 | VLDL cholesterol |
| L-LDL-L | l_ldl_l | HDL cholesterol |
| IDL-CE | idl_ce | Total triglycerides |
| M-HDL-CE | m_hdl_ce | Triglycerides in VLDL |
| M-LDL-PL | m_ldl_pl | Total esterified cholesterol |
| M-HDL-C | m_hdl_c | Average diameter for VLDL particles |
| M-HDL-L | m_hdl_l | Average diameter for HDL particles |
| M-HDL-PL | m_hdl_pl | Ratio of triglycerides to phosphoglycerides |
| MUFA | mufa | Apolipoprotein B |
| IDL-FC | idl_fc | Apolipoprotein A1 |
| L-LDL-TG % | l_ldl_tg_percentage | Ratio of apolipoprotein B to apolipoprotein A1 |
| IDL-C | idl_c | Total fatty acids |
| XS-VLDL-P | xs_vldl_p | Monounsaturated fatty acids |
| XL-VLDL-C | xl_vldl_c | Alanine |
| L-LDL-PL | l_ldl_pl | Glutamine |

| | | |
|---|---|---|
| Total fatty acids | totfa | Isoleucine |
| Phenylalanine | phe | Leucine |
| XL-VLDL-FC | xl_vldl_fc | Valine |
| IDL-L | idl_l | Phenylalanine |
| L-HDL-FC | l_hdl_fc | Tyrosine |
| L-LDL-P | l_ldl_p | Glucose |
| L-VLDL-CE | l_vldl_ce | Lactate |
| Total-CE | estc | Pyruvate |
| S-VLDL-CE | s_vldl_ce | Acetate |
| IDL-TG % | idl_tg_percentage | Creatinine |
| S-HDL-TG % | s_hdl_tg_percentage | Glycoprotein acetyls |
| XL-VLDL-PL | xl_vldl_pl | Concentration of very large VLDL particles |
| Total-C | serum_c | Total lipids in very large VLDL |
| TG/PG | tg_pg | Phospholipids in very large VLDL |
| Valine | val | Cholesterol in very large VLDL |
| L-HDL-CE | l_hdl_ce | Free cholesterol in very large VLDL |
| Acetate | ace | Triglycerides in very large VLDL |
| IDL-P | idl_p | Concentration of large VLDL particles |
| VLDL particle size | vldl_d | Total lipids in large VLDL |

BBMRI Consortium Banner

BBMRI.nl
Biobanking and BioMolecular resources Research Infrastructure The Netherlands

METABOLOMICS CONSORTIUM

#### COHORT COLLECTION:

J.M. Geleijnse[1], E. Boersma[2], W.E. van Spil[3], M.M.J. van Greevenbroek[4, 5], C.D.A. Stehouwer[4, 5], C.J.H. van der Kallen[4, 5], I.C.W. Arts[5, 6, 7], F. Rutters[8, 9], J.W.J. Beulens[8, 9], M. Muilwijk[8, 10], P.J.M. Elders[8, 10], L.M. 't Hart[8, 9, 11, 12], M. Ghanbari[13, 14], M.A. Ikram[13], M.G. Netea[15], M. Kloppenburg[16, 17], Y.F.M. Ramos[18], N. Bomer[19], I. Meulenbelt[18], K. Stronks[20], M.B. Snijder[20], A.H. Zwinderman[21], B.T. Heijmans[18], L.H.Lumey[22], C. Wijmenga[23], J. Fu[23, 24], A. Zhernakova[23], J. Deelen[25, 18], S.P. Mooijaart[26], M. Beekman[18], P.E. Slagboom[18, 25], G.L.J. Onderwater[27], A.M.J.M. van den Maagdenberg[28, 27], G.M. Terwindt[27], C.Thesing[29, 8], M. Bot[29, 8], B.W.J.H. Penninx[29, 8], S. Trompet[30, 26], J.W. Jukema[30], N. Sattar[31], I.C.C. van der Horst[32], P. van der Harst[33], C. So-Osman[34, 35], J.A. van Hilten[36], R.G.H.H. Nelissen[37], I.E. Höfer[38], F.W. Asselbergs[39, 40], P. Scheltens[41], C.E. Teunissen[42], W.M. van der Flier[43, 41], J. van Dongen[29, 8], R. Pool[29], A.H.M. Willemsen[29, 8], D.I. Boomsma[29, 8]

#### SAMPLE LOGISTICS, DATABASE & CATALOGUE:

H.E.D. Suchiman[18], J.J.H. Barkey Wolf[18], M. Beekman[18], D. Cats[45], H. Mei[45], M. Slofstra[23], M. Swertz[46, 23], M.J.T. Reinders[47, 48], E.B. van den Akker[47, 18]

#### STEERING COMMITTEE:

D.I. Boomsma[29, 8], M.A. Ikram[13], P.E. Slagboom[18, 25]

#### AFFILIATIONS:

1. Division of Human Nutrition and Health, Wageningen University, Wageningen, The Netherlands
2. Thorax centre, Erasmus Medical Centre, Rotterdam, the Netherlands
3. Department of Rheumatology & Clinical Immunology, University Medical Center Utrecht, Utrecht, The Netherlands
4. Department of Internal Medicine, Maastricht University Medical Center (MUMC+), Maastricht, The Netherlands
5. School for Cardiovascular Diseases (CARIM), Maastricht University, Maastricht, the Netherlands
6. Department of Epidemiology, Maastricht University, Maastricht, the Netherlands
7. Maastricht Center for Systems Biology, Maastricht University, Maastricht, the Netherlands
8. Amsterdam Public Health Research Institute, Amsterdam, The Netherlands
9. Department of Epidemiology and Biostatistics, Amsterdam University Medical Center, Vrije Universiteit, Amsterdam, the Netherlands
10. Department of General Practice and Elderly Care Medicine, Amsterdam University Medical Center, Vrije Universiteit, Amsterdam, the Netherlands
11. Department of Epidemiology and Biostatistics, Amsterdam University Medical Center, Vrije Universiteit, Amsterdam, the Netherlands
12. Department of Cell and Chemical Biology, Leiden University Medical Center, Leiden, the Netherlands
13. Department of Epidemiology, Erasmus MC, University Medical Center, Rotterdam, The Netherlands
14. Department of Genetics, School of Medicine,, Mashhad University of Medical Sciences, Mashhad, Iran
15. Department of Internal Medicine and Radboud Center for Infectious Diseases, Radboud University Medical Center, Nijmegen, The Netherlands

16. Department of Clinical Epidemiology, Leiden University Medical Centre, Leiden, The Netherlands
17. Department of Rheumatology, Leiden University Medical Center, The Netherlands
18. Department of Biomedical Data Sciences, Section of Molecular Epidemiology, Leiden University Medical Center, Leiden, The Netherlands
19. Department of Experimental Cardiology, University of Groningen, University Medical Center Groningen, Groningen, The Netherlands
20. Department of Public Health, Academic Medical Center, University of Amsterdam, Amsterdam, The Netherlands
21. Department of Clinical Epidemiology, Biostatistics, and Bioinformatics, Academic Medical Centre, University of Amsterdam, Amsterdam, The Netherlands
22. Department of Epidemiology, Mailman School of Public Health, Columbia University, New York, NY 10032
23. Department of Genetics, University Medical Center Groningen, Groningen, The Netherlands
24. Department of Pediatrics, University Medical Center Groningen, Groningen, The Netherlands
25. Max Planck Institute for Biology of Ageing, Cologne, Germany
26. Department of Internal Medicine, Division of Gerontology and Geriatrics, Leiden University Medical Centre, Leiden, The Netherlands
27. Department of Neurology, Leiden University Medical Center, Leiden, The Netherlands
28. Department of Human Genetics, Leiden University Medical Center, Leiden, The Netherlands
29. Department of Biological Psychology, Amsterdam University Medical Center, Vrije Universiteit, Amsterdam, The Netherlands
30. Department of Cardiology, Leiden University Medical Center, Leiden, The Netherlands
31. Institute of Cardiovascular and Medical Sciences, Cardiovascular Research Centre, University of Glasgow, Glasgow, UK
32. Department of Critical Care, University Medical Center Groningen, Groningen, The Netherlands
33. Department of Cardiology, University Medical Center Utrecht, Utrecht, The Netherlands
34. Sanquin Blood Bank, Leiden and Department of Haematology, Groene Hart Hospital, Gouda, the Netherlands
35. International Society of Blood Transfusion (ISBT), Amsterdam, The Netherlands
36. Unit of Transfusion Medicine, Sanquin Blood Bank, Leiden, The Netherlands
37. Department of Orthopaedics, Leiden University Medical Center, Leiden, The Netherlands
38. Department of Clinical Chemistry and Hematology, UMC Utrecht, the Netherlands
39. Department of Cardiology, Division Heart and Lungs, University Medical Center Utrecht, Utrecht, The Netherlands
40. Julius Center for Health Sciences and Primary Care, University Medical Center Utrecht, Utrecht, The Netherlands
41. Department of Neurology & Alzheimer Center, VU University Medical Center, Amsterdam, The Netherlands
42. Neurochemistry Laboratory, Clinical Chemistry Department, Amsterdam University Medical Center, Amsterdam Neuroscience, The Netherlands
43. Department of Epidemiology and Biostatistics, VU University Medical Center, Amsterdam, The Netherlands
44. SURFsara, Amsterdam, the Netherlands
45. Sequence Analysis Support Core, Leiden University Medical Center, Leiden, the Netherlands
46. University of Groningen, University Medical Center Groningen, Genomics Coordination Center, Groningen, the Netherlands
47. Leiden Computational Biology Center, Leiden University Medical Center, Leiden, the Netherlands
48. The Delft Bioinformatics Lab, Delft University of Technology, Delft, the Netherlands

#### Cohort Description

##### Alpha Omega Cohort

The Alpha Omega Cohort consists of 4,837 Dutch men and women aged 60-80 years with a clinically diagnosed myocardial infarction <10 years before study enrollment. Baseline examinations and blood sampling took place between 2002 and 2006, after which the cohort has been followed up for cause-specific mortality. During the first 3 years of follow-up, patients participated in an intervention study with omega-3 fatty acids (Alpha Omega Trial) [1,2]. For the present analysis, a random sample of 600 patients was selected, who were followed up through Statistics Netherlands (CBS) until the 1st of January 2014. Metabolites were successfully quantified in EDTA plasma samples of 568 patients. The Alpha Omega Cohort is registered with clinicaltrials.gov (Identifier: NCT03192410).

1. Geleijnse JM, Giltay EJ, Schouten EG, et al. Effect of low doses of n-3 fatty acids on cardiovascular diseases in 4,837 post-myocardial infarction patients: design and baseline characteristics of the Alpha Omega Trial. Am Heart J 2010;159:539-46 e2.
2. Kromhout D, Giltay EJ, Geleijnse JM, Alpha Omega Trial G. n-3 fatty acids and cardiovascular events after myocardial infarction. N Engl J Med 2010;363:2015-26.

##### Amsterdam Dementia Cohort

The Amsterdam Dementia Cohort is an ongoing study including patients who visit the memory clinic of the Alzheimer center of the VU University Medical Center [1]. At baseline, all subjects receive a diagnostic assessment including medical history taking, physical and neurological examination, neuropsychological investigation, standard laboratory tests of blood and cerebrospinal fluid, electroencephalogram and brain magnetic resonance imaging. Clinical diagnosis is made in consensus based, multidisciplinary meetings. For the present metabolomics analysis, 1,473 plasma samples were selected based on available volume in our biobank. In this cohort 45% is female and mean±SE age of 64±9. Date of enrollment was between 2001-2014.

1. van der Flier, W.M., et al., Optimizing patient care and research: the Amsterdam Dementia Cohort. J Alzheimers Dis, 2014. 41(1): p. 313-27.

##### BIOMARCS

The BIOMarker study to identify the Acute risk of a Coronary Syndrome (BIOMArCS) was designed to study the relation between temporal changes in cardiovascular biomarkers and ischemic cardiovascular events in patients discharged after acute coronary syndrome (ACS) admission.[1] 844 ACS patients were enrolled in 18 hospitals in The Netherlands. Venipuncture was scheduled at 19 regular intervals during a year. 45 Patients (cases) reached the study endpoint of repeat ACS within one year. BIOMArCS was approved by the institutional review committees of the participating hospitals. All patients gave informed consent.

1. Rohit M Oemrawsingh et al. , Cohort profile of BIOMArCS: the BIOMarker study to identify the Acute risk of a Coronary Syndrome-a prospective multicentre biomarker study conducted in the Netherlands; BMJ Open. 2016 Dec 23;6(12):e012929. doi: 10.1136/bmjopen-2016-012929.

##### LUMINA

The Leiden University MIgraine Neuro-Analysis (LUMINA) cohort currently consists of over 6,700 male and female participants aged 18-88 years. LUMINA participants are recruited through a dedicated, nationwide website (http://www.lumc.nl/org/hoofdpijn-onderzoek/onderzoek) inviting Dutch migraine patients and non-migraine controls to participate in migraine research. Participants recruited through the website were asked to complete a screening questionnaire, previously validated to diagnose migraine [1]. The questionnaire was validated by a semi-structured telephone interview, in accordance with the International Classification of Headache Disorders (ICDH-3β)[2]. In addition, patients attending the Leiden University Medical Centre (LUMC) dedicated headache clinic were invited to participate. Migraine patients recruited through the headache clinic were diagnosed with migraine by a neurologist specialized in headache. For the present metabolomics analysis, a sample of migraine patients and non-migraine controls was selected, who were participating in several LUMINA sub cohorts (CSF, MRS, and CHARM cohorts) between 2008-2014. Migraine diagnosis was confirmed by the study-physician at the day of blood draw resulting in the collection of 564 blood (EDTA/Serum) samples from 432 individual participants used for metabolite quantification.

1. van Oosterhout WPJ, Weller CM, Stam AH, et al. Validation of the web-based LUMINA questionnaire for recruiting large cohorts of migraineurs. Cephalalgia. 2011;31:1359–1367. doi: 10.1177/0333102411418846.
2. Headache Classification Committee of the International Headache Society (IHS). The International Classification of Headache Disorders, 3rd edition (beta version). Cephalalgia. 2013;33:629–808. doi: 10.1177/0333102413485658.

#### CHECK

CHECK (Cohort Hip & Cohort Knee) is a prospective, 10-year follow-up, observational cohort study of 1002 people aged between 45 and 65 years at the time of inclusion, with pain in their knee(s) and/or hip(s), who had never or not longer than 6 months ago consulted a physician for these complaints [1]. Blood samples were taken non-fasted. Hip and knee radiographs were obtained multiple times throughout follow-up and scored pairwise according to the Kellgren & Lawrence (KL) scoring system. When scored pairwise, these people did not have obvious radiographic knee or hip OA at baseline (i.e. KL0 or 1).

1. Wesseling J, Dekker J, van den Berg WB, Bierma-Zeinstra SM, Boers M, Cats HA, et al. CHECK (Cohort Hip and Cohort Knee): similarities and differences with the Osteoarthritis Initiative. Ann Rheum Dis 2009;68:1413-9.

#### CODAM

The Cohort on Diabetes and Atherosclerosis Maastricht [1] (CODAM) is a prospective, observational study that consists of 574 individuals who were selected from a larger population-based cohort [2]. Inclusion of participants into CODAM was based on a moderately increased risk to develop cardiometabolic diseases, such as type 2 diabetes and/or cardiovascular disease. Participants were included if they were of Caucasian descent and over 40 yrs of age and additionally met at least one of the following criteria: increased BMI (>25 kg/m$^2$), a positive family history of type 2 diabetes, a history of gestational diabetes and/or glycosuria, or use of anti-hypertensive medication. The CODAM baseline measurements were done between 2000 and 2002 (n=352 men, 222 women, age 59.6±7.0 yrs). After a median of 7.0 years (interquartile range 6.9–7.1), 495 participants were included in the follow-up measurements [3]. Metabolites were obtained fasting EDTA plasma samples of all participants at baseline and at follow-up in those participants who had type 2 diabetes at baseline (n=110).

1. van Greevenbroek, M. M. J. et al. The cross-sectional association between insulin resistance and circulating complement C3 is partly explained by plasma alanine aminotransferase, independent of central obesity and general inflammation (the CODAM study). Eur. J. Clin. Invest. 41, 372–379 (2011). doi: 10.1111/j.1365-2362.2010.02418.x
2. Van Dam, R. M., Boer, J. M., Feskens, E. J. M. & Seidell, J. C. Parental history of diabetes modifies the association between abdominal adiposity and hyperglycemia. Diabetes Care 24, 1454–1459 (2001).
3. Wlazlo N, van Greevenbroek MM, Ferreira I, Feskens EJ, van der Kallen CJ, Schalkwijk CG, Bravenboer B, Stehouwer CD. Complement factor 3 is associated with insulin resistance and with incident type 2 diabetes over a 7-year follow-up period: the CODAM Study. Diabetes Care37, 1900-1909 (2014). doi: 10.2337/dc13-2804

#### The Maastricht Study (TMS)

The Maastricht Study [1] is an observational prospective population-based cohort study enriched with T2DM individuals. Eligible for participation were individuals aged between 40 and 75 years and living in the southern part of the Netherlands (municipalities Maastricht, Margraten-Eijsden, Meersen and Valkenburg; Maastricht and Heuvelland in the province of Limburg).

1. Schram MT1 et al. The Maastricht Study: an extensive phenotyping study on determinants of type 2 diabetes, its complications and its comorbidities. Eur J Epidemiol. 2014 Jun;29(6):439-51. doi: 10.1007/s10654-014-9889-0. Epub 2014 Apr 23.

#### The Hoorn Diabetes Care System cohort study (DCS)

The DCS cohort currently consists of approximately 13.000people with type 2 diabetes. The DCS provides routine diabetes care to people with type 2 diabetes living in the West-Friesland region of the Netherlands [1]. People treated by the DCS visit the DCS research center annually, during which blood is drawn in the fasting state for routine biochemistry. Furthermore, all participants get a full medical exam, advice about their health and treatment and receive education on their disease during their annual visits to the DCS research center. In addition, patients are invited to join our research and biobanking studies (n=5,000+). From the DCS biobank we included for this study a random cross-sectional sample for which a baseline plasma sample and yearly follow-up data were available (n=750). For case-control analyses this sample was supplemented with individuals selected for the inability to reach the glycaemic target (HbA1c>53 mmol/mol) and/or suffering from diabetic complications (n=245). Samples were collected in 2008/2009 and stored and -80 degrees Celsius until analysis. Metabolites were successfully quantified in fasting EDTA plasma samples from 995 individuals.

[1] van der Heijden AA, Rauh SP, Dekker JM, Beulens JW, Elders P, 't Hart LM, Rutters F, van Leeuwen N, Nijpels G: The Hoorn Diabetes Care System (DCS) cohort. A prospective cohort of persons with type 2 diabetes treated in primary care in the Netherlands. BMJ Open 2017;7:e015599

**Website**: www.hoornstudies.com

**Erasmus Rucphen Family study (ERF)**

The Erasmus Rucphen Family is a family-based study that includes inhabitants of a genetically isolated community in the South-West of the Netherlands [1]. The goal of the study is to identify the risk factors in the development of complex disorders. Study population includes approximately 3,000 individuals who are living descendants of a limited number of founders living in the 19th century [1]. Metabolomics measurements were quantified from fasted EDTA plasma samples using Nightingale Health platform. All data were collected between 2002 and 2005. Metabolomics measurements were available for 1,402 participants from the ERF.

1. Pardo, Luba M., et al. "The effect of genetic drift in a young genetically isolated population." Annals of human genetics69.3 (2005): 288-295.

**Rotterdam Study (RS)**

The Rotterdam Study is a prospective, population-based cohort study among individuals living in the well-defined Ommoord district in the city of Rotterdam in The Netherlands [1]. The aim of the study is to determine the occurrence of cardiovascular, neurological, ophthalmic, endocrine, hepatic, respiratory, locomotor, dermatological, otolaryngological, and psychiatric diseases in elderly people. The cohort was initially defined in 1990 among approximately 7,983 persons, aged 55 years and older, who underwent a home interview and extensive physical examination at the baseline and during follow-up visits every 3-4 years (RS-I)[1]. Cohort was extended in 2000/2001 (RS-II, 3,011 individuals aged 55 years and older) and 2006/2008 (RS-III, 3,932 subjects, aged 45 and older). As of 2008, Rotterdam Study comprised 14,926 subjects. Written informed consent was obtained from all participants and the Medical Ethics Committee of the Erasmus Medical Center, Rotterdam, approved the study. Metabolomics measurements were quantified in fasted EDTA plasma samples using Nightingale Health platform. Metabolomics measurements were available for 2,986 participants from RS-I, 591 participants from RS-II (n=591) and 1,787 participants from RS-III.

1. Ikram, M. Arfan, et al. "The Rotterdam Study: 2018 update on objectives, design and main results." *European Journal of Epidemiology* 32.9 (2017): 807-850.

**FUNCTGENOMICS**

The 500 Functional Genomics (500FG [1]) project consists of 534 adult healthy volunteers sampled between July 2013 and December 2014. Inclusion criteria were >18 years of age and Western European descent. Exclusion criteria were pregnancy/breastfeeding, chronic or acute disease at the time of assessment, and use of chronic or acute medication during the last month before the study. After visiting the hospital to donate blood, volunteers received an extensive online questionnaire about lifestyle, diet, and disease history. Upon analyzing the questionnaire data we excluded 45 volunteers as they were under medication, non-European descent, or had kidney disease or diabetes mellitus.

1. Schirmer et al. Linking the Human Gut Microbiome to Inflammatory Cytokine Production Capacity. Cell. 2016 Nov 3;167(4):1125-1136.e8. doi: 10.1016/j.cell.2016.10.020.

GARP

The GARP cohort (N=217) consists of patients with advanced radiographic OA at two or more joint sites of hand, spine, knee or hip. Follow-up was performed at 5 years, at which radiographs for hip, knee and hand were scored pairwise using the OARSI Atlas and the KL scoring system. Matched to the GARP study, a normal reference control group (NORREF) was collected using the same protocol and included in this study as controls [1-3]. Blood was collected non-fasted.

1. Riyazi N, Meulenbelt I, Kroon HM, Ronday KH, Hellio le Graverand MP, Rosendaal FR, et al. Evidence for familial aggregation of hand, hip, and spine but not knee osteoarthritis in siblings with multiple joint involvement: the GARP study. Ann Rheum Dis 2005;64:438-43.

2. Meulenbelt I, Kloppenburg M, Kroon HM, Houwing-Duistermaat JJ, Garnero P, Hellio-Le Graverand MP, et al. Clusters of biochemical markers are associated with radiographic subtypes of osteoarthritis (OA) in subject with familial OA at multiple sites. The GARP study. Osteoarthritis Cartilage 2007;15:379-85.

3. Bijsterbosch J, Meulenbelt I, Watt I, Rosendaal FR, Huizinga TW, Kloppenburg M. Clustering of hand osteoarthritis progression and its relationship to progression of osteoarthritis at the knee. Ann Rheum Dis 2014;73:567-72.

HELIUS study

The HELIUS study is a prospective cohort study among the largest ethnic groups living in Amsterdam, the Netherlands. The aim of the HELIUS study is to investigate the causes of (the unequal burden of) diseases across ethnic groups, focusing on three disease categories: cardiovascular diseases, mental health and infectious diseases [1]. Between 2011-2015, a total 24,789 participants (men and women aged 18-70 years) were included at baseline. Similar-sized samples of individuals of Dutch, African Surinamese, South-Asian Surinamese, Ghanaian, Turkish and Moroccan origin were included. Participants filled in an extensive questionnaire and underwent a physical examination that included the collection of biological samples (biobank). Follow-up data is obtained by linkages with existing registries (e.g. hospital data, insurance data) and will be obtained by repeated measurements [2]. Metabolites were quantified in EDTA plasma samples of 500 African origin participants with (pre)diabetes (235 African Surinamese, 265 Ghanaian participants).

[1] K Stronks, MB Snijder, RJ Peters, M Prins, AH Schene, AH Zwinderman. Unravelling the impact of ethnicity on health in Europe: the HELIUS study. BMC Public Health 2013;13:402.

[2] MB Snijder, H Galenkamp, M Prins, EM Derks, RJ Peters, AH Zwinderman, K Stronks. Cohort Profile: the Healthy Life in an Urban Setting (HELIUS) study. BMJ Open (in press).

**Website:** www.heliusstudie.nl

LIFELINES-DEEP

The LifeLines-DEEP cohort is a subset of the Dutch general population cohort LifeLines. Both LifeLines and LifeLines DEEP have been previously described [1-3]. In summary, LifeLines is a three-generation observational follow-up study, which was set up to investigate universal risk factors and their modifiers for multifactorial diseases. Since 2006, approximately 167,000 individuals from the general population residing in the three northern provinces of the Netherlands participate in the study. All participants will be followed-up prospectively for at least 30 years. Participants regularly undergo physical examinations and fill in extensive questionnaires. In addition, blood and urine samples are collected. Each participant is asked to fill in health, lifestyle, and quality-of-life questionnaires every 1.5 years, whereas each participant is invited for a follow-up visit to a Lifelines clinic every 5 years [1,2]. LifeLines-DEEP comprises 1,539 participants (636 males and 903 females, age range 18–84 years). This study was set up for the more detailed phenotyping and omics profiling. For analysis of the genome, epigenome, transcriptome, microbiome, metabolome and other biological levels, additional biomaterials were collected, including additional blood, exhaled air and fecal samples, as well as responses to gastrointestinal health [3].

The current metabolomics study included the baseline information and plasma samples of LifeLines-DEEP participants. EDTA plasma samples were collected after overnight fasting. Peripheral blood samples were drawn by venipuncture from the median cubital vein and subsequently placed at 4°C. Transport of the samples from the research site to the LifeLines laboratory in Groningen was under tightly controlled and continuously monitored conditions. At the LifeLines site, plasma was prepared and aliquoted and stored at -80°C. The samples underwent two freeze-thaw cycles prior to shipment to Brainshake for metabolome analysis [1]. With some sample drop-off, this study eventually included 1,440 LifeLines-DEEP participants. The LifeLines DEEP study was approved by the institutional ethics review board of University Medical Center Groningen (ref. M12.113965).

1. Scholtens, S. et al. Cohort Profile: LifeLines, a three-generation cohort study and biobank. Int. J. Epidemiol. 44, 1172–1180 (2015).

2. Stolk, R. P. et al. Universal risk factors for multifactorial diseases: LifeLines: A three-generation population-based study. Eur. J. Epidemiol. 23, 67–74 (2008).

3. Tigchelaar E. F. et al., Cohort profile: LifeLines DEEP, a prospective, general population cohort study in the northern Netherlands: study design and baseline characteristics. BMJ Open 5, e006772 (2015).

**Website:** https://www.lifelines.nl/

**Leiden Longevity Study (LLS)**

The Leiden Longevity Study (LLS) consists of 421 long-lived families of European descent. Families were included if at least two long-lived siblings were alive and fulfilled the age criterion of 89 years or older for males and 91 years or older for females, representing <0·5% of the Dutch population in 2001 [1]. In total, 944 long-lived proband siblings (mean age = 94 years, range = 89-104), 1671 offspring (mean age = 61 years, range = 39-81) and 744 spouses thereof (mean age = 60 years, range = 36-79) were included. Registry-based follow-up until the 27th of October 2016 was available for all participants. Metabolites were successfully quantified in 843 nonagenarians [LLS_SIBS], 1157 of their offspring and 684 controls (LLS_PAROFF) using non-fasted EDTA plasma samples.

1. M. Schoenmaker et al. Evidence of genetic enrichment for exceptional survival using a family approach: the Leiden Longevity Study. Eur J Hum Genet. 2006 Jan;14(1):79-84.

**NESDA**

The Netherlands Study of Depression and Anxiety (NESDA) is an ongoing observational longitudinal cohort study on the long term course and consequences of depressive and anxiety disorders [1]. Between September 2004 and February 2007, 2,981 participants (1,979 females, 1,002 males) aged 18 through 65 years were included. They were recruited through different settings (community, primary care and specialized mental health clinics) in order to obtain a representative sample of persons with depressive and/or anxiety disorders (in lifetime, n=2,329) and without depressive and/or anxiety disorders (n=652). At baseline, participants completed the 4-hour baseline assessment, which included a face-to-face interview, written questionnaires, and biological measurements. Follow-up visits to the research center have now been finished 2, 4, 6 and 9 years after baseline, with response rates of n=2,596, n=2,402, n=2,256 and n=2,069, respectively. The research protocol was approved by the Ethical Committee of the participating centers, and all participants provided written informed consent. During the baseline interview, EDTA plasma samples were collected and stored in aliquots at -85°C until further analysis. Participants were instructed to have an overnight fast before blood collection. Metabolites in these blood samples were analyzed in 2 batches (April and December 2014, respectively) by Brainshake Ltd./Nightingale Health, Helsinki, Finland.

[1] Penninx BWJH, Beekman ATF, Smit JH, Zitman FG, Nolen WA, Spinhoven P, et al. The Netherlands Study of Depression and Anxiety (NESDA): rationale, objectives and methods. Int J Methods Psychiatr Res 2008;17:121–40. doi: 10.1002/mpr.256.

**Website:** http://www.nesda.nl

**PROspective Study of Pravastatin in the Elderly at Risk (PROSPER)**

The PROspective Study of Pravastatin in the Elderly at Risk (PROSPER) trial design has been published [1,2]. In brief, 5,804 elderly adults (70-82 years old) were enrolled. This was a double-blind, randomised placebo controlled trial investigating the benefit of pravastatin (40 mg/day) in elderly individuals at risk of CVD. Participants were identified in the primary care setting from 3 centres: Glasgow, Scotland; Cork, Ireland or Leiden, the Netherlands. All participants had high-normal to high cholesterol (4·0-9·0 mmol/L) at baseline. Additionally 50% of patients had evidence of vascular disease (physician diagnosed stable angina, stroke, transient ischaemic attack (TIA) or myocardial infarction (MI)) and the remaining 50% of patients had high risk of vascular disease as they had either hypertension, diabetes or were smokers. The primary outcome measure of PROSPER was a composite CVD outcome. In the current study the endpoint of interest was all-cause mortality. Patients were recruited between December 1997 and May 1999 and the mean follow-up period was 3·2 years. Fasting venous blood samples were collected at baseline and at 3-month intervals and biobanked at -80°C. For the present study previously unthawed 6-month post-randomisation samples were used, employing the study as a cohort study and adjusting for randomised treatment in the analyses. Metabolites were successfully quantified in 5,329 individuals.

1. Shepherd J, Blauw GJ, Murphy MB, Cobbe SM, Bollen EL, Buckley BM, et al. The design of a prospective study of Pravastatin in the Elderly at Risk (PROSPER). PROSPER Study Group. PROspective Study of Pravastatin in the Elderly at Risk. Am J Cardiol 1999 Nov 15;84(10):1192-7. PMID: 10569329

2. Shepherd J, Blauw GJ, Murphy MB, Bollen EL, Buckley BM, Cobbe SM, et al. Pravastatin in elderly individuals at risk of vascular disease (PROSPER): a randomised controlled trial. Lancet 2002 Nov 23;360(9346):1623-30. PMID:12457784

**LUMC Arthroplasty studies:**

The LUMC arthroplasty studies (N=462) consist of participants of the RAAK, TacTics (NTR309) and TOMaat (NTR303) studies [1, 2]. These cross-sectional studies included OA patients who received THA or TKA. Since all participants underwent THA/TKA, all patients are considered as end-stage OA and included in the cross-sectional OA prevalence analysis. Blood samples were collected during surgery while patients were fasted.

[1] Ramos YF, den Hollander W, Bovee JV, Bomer N, van der Breggen R, Lakenberg N, et al. Genes involved in the osteoarthritis process identified through genome wide expression analysis in articular cartilage; the RAAK study. PLoS One 2014;9:e103056.

[2] So-Osman C, Nelissen RG, Koopman-van Gemert AW, Kluyver E, Poll RG, Onstenk R, et al. Patient blood management in elective total hip- and knee-replacement surgery (Part 1): a randomized controlled trial on erythropoietin and blood salvage as transfusion alternatives using a restrictive transfusion policy in erythropoietin-eligible patients. Anesthesiology 2014;120:839-51.

**STEMI-GIPS-III**

The Glycometabolic Intervention as Adjunct to Primary Coronary Intervention in ST Elevation Myocardial Infarction (GIPS-III) study is a placebo-controlled randomized clinical trial to evaluate the effect of metformin therapy on left ventricular function in 380 non-diabetic ST-elevated myocardial infarction (STEMI) patients aged 23-90. Patients were included between 2011 and 2013. Blood samples were collected at several time points after inclusion. During the 4-month treatment period, patients received metformin or placebo. 4 months after randomization, left ventricular ejection fraction (LVEF) was measured by cardiac MRI. In addition, patients were followed up for major adverse cardiac events (death, recurrent MI, target lesion revascularization), stroke, non-elective hospitalizations for chest pain or heart failure, all recurrent coronary interventions and internal cardiac defibrillator implantations. Metabolic profiling was assessed in EDTA plasma samples collected at baseline (hospital admission), 24 h post-MI and 4 months post-MI. GIPS-III is registered with clinicaltrials.gov (ID: NCT01217307)

1. Eppinga RN, Kofink D, Dullaart RP, Dalmeijer GW, Lipsic E, van Veldhuisen DJ, van der Horst IC, Asselbergs FW, van der Harst P. Effect of Metformin on Metabolites and Relation With Myocardial Infarct Size and Left Ventricular Ejection Fraction After Myocardial Infarction. Circ Cardiovasc Genet. 2017 Feb;10(1).

2. Eppinga RN, Hartman MH, van Veldhuisen DJ, Lexis CP, Connelly MA, Lipsic E, van der Horst IC, van der Harst P, Dullaart RP. Effect of Metformin Treatment on Lipoprotein Subfractions in Non-Diabetic Patients with Acute Myocardial Infarction: A Glycometabolic Intervention as Adjunct to Primary Coronary Intervention in ST Elevation Myocardial Infarction (GIPS-III) Trial. PLoS One. 2016 Jan 25;11(1):e0145719.

3. Lexis CP, van der Horst-Schrivers AN, Lipsic E, Valente MA, Muller Kobold AC, de Boer RA, van Veldhuisen DJ, van der Harst P, van der Horst IC. The effect of metformin on cardiovascular risk profile in patients without diabetes

presenting with acute myocardial infarction: data from the Glycometabolic Intervention as adjunct to Primary Coronary Intervention in ST Elevation Myocardial Infarction (GIPS-III) trial. BMJ Open Diabetes Res Care. 2015 Dec 11;3(1):e000090.

4. Lexis CP, van der Horst IC, Lipsic E, Wieringa WG, de Boer RA, van den Heuvel AF, van der Werf HW, Schurer RA, Pundziute G, Tan ES, Nieuwland W, Willemsen HM, Dorhout B, Molmans BH, van der Horst-Schrivers AN, Wolffenbuttel BH, ter Horst GJ, van Rossum AC, Tijssen JG, Hillege HL, de Smet BJ, van der Harst P, van Veldhuisen DJ; GIPS-III Investigators. Effect of metformin on left ventricular function after acute myocardial infarction in patients without diabetes: the GIPS-III randomized clinical trial. JAMA. 2014 Apr 16;311(15):1526-35.

5. Lexis CP, van der Horst IC, Lipsic E, van der Harst P, van der Horst-Schrivers AN, Wolffenbuttel BH, de Boer RA, van Rossum AC, van Veldhuisen DJ, de Smet BJ; GIPS-III Investigators. Metformin in non-diabetic patients presenting with ST elevation myocardial infarction: rationale and design of the glycometabolic intervention as adjunct to primary percutaneous intervention in ST elevation myocardial infarction (GIPS)-III trial. Cardiovasc Drugs Ther.

UCORBIO

The Utrecht Coronary Biobank Study (UCORBIO) enrolled 2,591 patients aged 18-93 who underwent coronary angiography for any indication at the UMC Utrecht. Baseline assessment and blood sampling took place between 2011 and 2014. Patients were followed up for the occurrence of major adverse cardiovascular events (stroke, myocardial infarction, coronary revascularization, death). During the follow-up period (maximum: 3 years), patients completed questionnaires every year to obtain information on hospital admissions. General practitioners and hospitals were contacted to confirm reported cardiovascular events. EDTA samples of 1,198 patients were selected for metabolic profiling. UCORBIO is registered with clinicaltrials.gov (ID: NCT02304744).

1. Gijsberts CM, Santema BT, Asselbergs FW, de Kleijn DP, Voskuil M, Agostoni P, Cramer MJ, Vaartjes I, Hoefer IE, Pasterkamp G, den Ruijter HM. Women Undergoing Coronary Angiography for Myocardial Infarction or Who Present With Multivessel Disease Have a Poorer Prognosis Than Men. Angiology. 2016 Jul;67(6):571-81.

2. Gijsberts CM, Agostoni P, Hoefer IE, Asselbergs FW, Pasterkamp G, Nathoe H, Appelman YE, de Kleijn DP, den Ruijter HM. Gender differences in health-related quality of life in patients undergoing coronary angiography. Open Heart. 2015 Aug 27;2(1):e000231.

3. Gijsberts CM, Gohar A, Ellenbroek GH, Hoefer IE, de Kleijn DP, Asselbergs FW, Nathoe HM, Agostoni P, Rittersma SZ, Pasterkamp G, Appelman Y, den Ruijter HM. Severity of stable coronary artery disease and its biomarkers differ between men and women undergoing angiography. Atherosclerosis. 2015 Jul;241(1):234-40.

VUNTR

Since 1987, the Netherlands Twin Register is collecting (longitudinal) data in young and adult twins and their families [1,2]. The rich phenotypic longitudinal information that has been collected extends from lifestyle, exposures, personality and demographics information to mental and somatic health. In subgroups information on autonomic and central nervous system function, biomarkers and gene expression, epigenetics and genotyping is available. A 2015 estimate is that, since initiating the NTR, ~25% of all twins and multiples in the Netherlands participated in NTR research projects. Longitudinal information for over 200,000 participants (twins, multiples and family members) was collected over multiple NTR research projects. Data collection is ongoing. A pdf of nearly all published papers may be found at the NTR website.

As part of a Netherlands Twin Register (NTR) biobank project (BB1), 9,530 participants from 3,477 families were visited at home between January 2004 and July 2008 for collection of blood samples [3]. A second project (BB2) collected blood samples in 517 subjects from January 2011 to December 2011, including 210 MZ twin pairs and 64 twin-spouse pairs [4]. Visits were scheduled between 7:00 and 10:00 am and fertile women were bled on day 2-4 of the menstrual cycle, or in their pill-free week. Body composition was measured and information about physical health and lifestyle (e.g. smoking and drinking behavior, exercise, medication use) was obtained. For more detailed information about the methodology of the NTR Biobank study, see [3]. The NTR studies were approved by the Central Ethics Committee on Research involving human subjects of the VUMC, Amsterdam, an Institutional Review Board certified by the US Office of Human Research Protections (IRB number IRB-2991 under Federal wide Assurance-3703; IRB/institute codes, NTR 03-180). All subjects provided written informed consent. Subject were selected who were part of NTR biobank and who in general had rich phenotyping data available.

1. Van Beijsterveldt CE, Groen-Blokhuis M, Hottenga JJ, Franić S, Hudziak JJ, Lamb D, Huppertz C, de Zeeuw E, Nivard M, Schutte N, Swagerman S, Glasner T, van Fulpen M, Brouwer C, Stroet T, Nowotny D, Ehli EA, Davies GE, Scheet P, Orlebeke JF, Kan KJ, Smit D, Dolan CV, Middeldorp CM, de Geus EJ, Bartels M, Boomsma DI. The Young Netherlands Twin Register (YNTR): longitudinal twin and family studies in over 70,000 children. *Twin Res Hum Genet.* **2013** Feb; 16(1): 252-267. doi: 10.1017/thg.2012.118. PMID: 23186620

2. Willemsen G, Vink JM, Abdellaoui A, den Braber A, van Beek JH, Draisma HH, van Dongen J, van 't Ent D, Geels LM, van Lien R, Ligthart L, Kattenberg M, Mbarek H, de Moor MH, Neijts M, Pool R, Stroo N, Kluft C, Suchiman HE, Slagboom PE, de Geus EJ, Boomsma DI.The Adult Netherlands Twin Register: twenty-five years of survey and biological data collection.*Twin Res Hum Genet.* **2013** Feb; 16(1): 271-281. doi: 10.1017/thg.2012.140. PMID: 23298648. PMCID: PMC3739974.

3. Willemsen G, de Geus EJ, Bartels M, van Beijsterveldt CE, Brooks AI, Estourgie-van Burk GF, Fugman DA, Hoekstra C, Hottenga JJ, Kluft K, Meijer P, Montgomery GW, Rizzu P, Sondervan D, Smit AB, Spijker S, Suchiman HE, Tischfield JA, Lehner T, Slagboom PE, Boomsma DI. The Netherlands Twin Register biobank: a resource for genetic epidemiological studies. *Twin Res Hum Genet.* **2010** Jun; 13(3): 231-245. doi: 10.1375/twin.13.3.231. PMID: 20477721.

4. Sirota M, Willemsen G, Sundar P, Pitts SJ, Potluri S, Prifti E, Kennedy S, Ehrlich SD, Neuteboom J, Kluft C, Malone KE, Cox DR, de Geus EJ, Boomsma DI. Effect of genome and environment on metabolic and inflammatory profiles. *PLoS One.* **2015** Apr 8; 10(4): e012089. doi: 10.1371/journal.pone.0120898. PMID: 25853885. PMCID: PMC4390246.

**website:** www.tweelingenregister.org/

HOF

The HOF study is based on the follow-up of selected births in 1943-1947 in the Amsterdam and Rotterdam Midwife training schools and the Leiden University Department of Obstetrics. The aim of the study was to assess the long-term health impact of pre-natal exposure to the Dutch famine of 1944–1945. The study includes pre-famine and post-famine births in these clinics as time controls and same-sex siblings as family controls. Selected births were traced through population registries in the Netherlands and underwent a telephone interview and extensive physical examinations in 2003-2005. They also provided blood samples and specific informed consents for further studies on risk factors for cardiovascular and metabolic disease. Metabolomic measurements were available for 971 men and women of the study.

1. Lumey LH, Stein AD, Kahn HS, van der Pal-de Bruin KM, Blauw GJ, Zybert PA, et al. Cohort profile: the Dutch Hunger Winter families study. International Journal of Epidemiology 2007; 36(6):1196-204.

STABILITEIT

The STABILITEIT cohort contains subsample of the Amsterdam Dementia Cohort study (ADC). The aim of the study was to study the effects of long-ter storage on the metabolic biomarkers, over a period of 14 years.

#### Acknowledgements

**Alpha Omega Cohort**

Financial support for the Alpha Omega Cohort was obtained from the Dutch Heart Foundation (grant 200T401) and the National Institutes of Health (grant R01HL076200), USA. DNA isolation was funded by BBMRI-NL (grant CP2011-18).

**Amsterdam Dementia Cohort**

Research within the VUmc Alzheimer center is part of the neurodegeneration research program of Amsterdam Neuroscience supported by Alzheimer Nederland and Stichting VUmc fonds.

**BIOMARCS**

BIOMArCS was funded by the Netherlands Heart Foundation (grant 2007B012), the Netherlands Heart Institute, the Working Group Cardiovascular Research Netherlands, and Eli Lilly through unrestricted research grants. The funders had no role in the design and conduct of the study; collection, management, analysis and interpretation of the data; preparation, review or approval of the manuscript; or decision to submit the manuscript for publication.

**LUMINA**

LUMINA (i.e. its subcohorts CHARM, CSF, MRS) is supported by grants obtained from the Netherlands Organization for the Health Research and Development (ZonMw; no. 90700217) and VIDI (ZonMw; no. 91711319); the Netherlands Organisation for Scientific Research (NWO) VICI (no.918.56.602) and Spinoza prize (2009) grants; the Centre for Medical Systems Biology (CMSB) and Netherlands Consortium for Systems Biology (NCSB), both within the framework of the Netherlands Genomics Initiative (NGI)/Netherlands Organization for Scientific Research (NWO) the 7th Framework EU project EUROHEADPAIN (no. 602633).

**CHECK**

CHECK was funded by the Dutch Arthritis Association on the lead of a steering committee comprising 16 members with expertise in different fields of OA, chaired by Professor JWJ Bijlsma and coordinated by J Wesseling. Involved are: Erasmus Medical Center Rotterdam; Kennemer Gasthuis Haarlem; Leiden University Medical Center; Maastricht University Medical Center; Martini Hospital Groningen/Allied Health Care Center for Rheumatology and Rehabilitation Groningen; Medical Spectrum Twente Enschede/Ziekenhuisgroep Twente Almelo; Reade, formerly Jan van Breemen Institute/VU Medical Center Amsterdam; St. Maartenskliniek Nijmegen; University Medical Center Utrecht, and Wilhelmina Hospital Assen.

**CODAM**

The initiation of the CODAM study was supported by grants of the Netherlands Organisation for Scientific Research (940-35-034) and the Dutch Diabetes Research Foundation (98·901).

**The Maastricht Study (TMS)**

This study was supported by the European Regional Development Fund via OP-Zuid, the Province of Limburg, the Dutch Ministry of Economic Affairs (grant 31O.041), Stichting De Weijerhorst (Maastricht, the Netherlands), the Pearl String Initiative Diabetes (Amsterdam, the Netherlands), CARIM School for Cardiovascular Diseases (Maastricht, the Netherlands), Stichting Annadal (Maastricht, the Netherlands), Health Foundation Limburg (Maastricht, the Netherlands) and by unrestricted grants from Janssen-Cilag B.V. (Tilburg, the Netherlands), Novo Nordisk Farma B.V. (Alphen aan den Rijn, the Netherlands) and Sanofi-Aventis Netherlands B.V. (Gouda, the Netherlands).

**The Hoorn Diabetes Care System cohort study (DCS)**

This study has been made possible by collaboration with the Diabetes Care System West-Friesland. The authors thank participants of this study and research staff of the Diabetes Care System West-Friesland.

**Erasmus Rucphen Family (ERF) study**

The Erasmus Rucphen Family (ERF) study has received funding from the Centre for Medical Systems Biology (CMSB) and Netherlands Consortium for Systems Biology (NCSB), both within the framework of the Netherlands Genomics Initiative (NGI)/Netherlands Organization for Scientific Research (NWO). ERF study is also a part of EUROSPAN (European Special Populations Research Network) (FP6 STREP grant number 18947 (LSHG-CT-2006-018947)); European Network of Genomic and Genetic Epidemiology (ENGAGE) from the European Community's Seventh Framework Programme (FP7/2007-2013)/grant agreement HEALTH-F4-2007-201413; "Quality of Life and Management of the Living Resources" of 5th Framework Programme (no. QLG2-CT-2002-01254); FP7 project EUROHEADPAIN (nr 602633), the Internationale Stichting Alzheimer Onderzoek (ISAO); the Hersenstichting Nederland (HSN); and the JNPD under the project PERADES (grant number 733051021, Defining Genetic, Polygenic and Environmental Risk for Alzheimer's Disease using multiple powerful cohorts, focused Epigenetics and Stem cell metabolomics). Metabolomics measurements of ERF has been funded by Biobanking and Biomolecular Resources Research Infrastructure (BBMRI)-NL (184.021.007). The ERF-follow up study is funded by CardioVasculair Onderzoek Nederland (CVON 2012-03). We are grateful to all study participants and their relatives, general practitioners and neurologists for their contributions and to P. Veraart for her help in genealogy, J. Vergeer for the supervision of the laboratory work, both S.J. van der Lee and A. van der Spek for collection of the follow-up data and P. Snijders for his help in data collection of both baseline and follow-up data.

**Rotterdam Study**

The Rotterdam Study is supported by the Erasmus MC University Medical Center and Erasmus University Rotterdam; The Netherlands Organisation for Scientific Research (NWO); The Netherlands Organisation for Health Research and Development (ZonMw); the Research Institute for Diseases in the Elderly (RIDE); The Netherlands Genomics Initiative (NGI); the Ministry of Education, Culture and Science; the Ministry of Health, Welfare and Sports; the European Commission (DG XII); and the Municipality of Rotterdam. The authors are grateful to the study participants, the staff from the Rotterdam Study and the participating general practitioners and pharmacists. Metabolomics measurements were funded by Biobanking and Biomolecular Resources Research Infrastructure (BBMRI)–NL (184.021.007) and the JNPD under the project PERADES (grant number 733051021, Defining Genetic, Polygenic and Environmental Risk for Alzheimer's Disease using multiple powerful cohorts, focused Epigenetics and Stem cell metabolomics).

**FUNCTGENOMICS**

**GARP**

The Leiden University Medical Centre have and are supporting the RAAK and GARP study. This study was supported by the Dutch Arthritis Foundation and Pfizer Groton, Connecticut, USA. We are indebted to drs. N. Riyazi, J. Bijsterbosch, H.M. Kroon and I. Watt for collection of data.

**HELIUS**

The HELIUS study is conducted by the Academic Medical Center Amsterdam and the Public Health Service of Amsterdam. Both organisations provided core support for HELIUS. The HELIUS study is also funded by the Dutch Heart Foundation, the Netherlands Organization for Health Research and Development (ZonMw), the European Union (FP-7), and the European Fund for the Integration of non-EU immigrants (EIF).

**LIFELINES-DEEP**

We thank the participants and staff of the Lifelines cohort for their collaboration, particularly B. Bolmer and S. Gerritsma for coordinating the Lifelines data. We thank J. Dekens and J. Arends for management and technical support and the Genomics Coordination Center for providing data infrastructure and access to high performance computing clusters. This project was funded by the Netherlands Heart Foundation (IN-CONTROL CVON grant 2012-03 and 2018-27), the grant from the Top Institute Food and Nitrition (TiFN GH001), the Netherlands Organization for Scientific Research (NWO) 024.004.017, 864.13.013, 016.178.056, 917.14.374, SPI 92-266; the European Research Council Consolidate grant 101001678; and the RuG Investment Agenda Grant Personalized Health grant.

**Leiden Longevity Study (LLS)**

The LLS has received funding from the European Union's Seventh Framework Programme (FP7/2007-2011) under grant agreement n° 259679. This study was supported by a grant from the Innovation-Oriented Research Program on Genomics (SenterNovem IGE05007), the Centre for Medical Systems Biology, and the Netherlands Consortium for Healthy Ageing (grants 05040202 and 050-060-810), all in the framework of the Netherlands Genomics Initiative, Netherlands Organization for Scientific Research (NWO), Unilever Colworth, and by BBMRI-NL, a Research Infrastructure financed by the Dutch government (NWO 184.021.007).

**NESDA**

The infrastructure for the NESDA study (www.nesda.nl) is funded through the Geestkracht program of the Netherlands Organisation for Health Research and Development (ZonMw, grant number 10-000-1002) and financial contributions by participating universities and mental health care organizations (VU University Medical Center, GGZ inGeest, Leiden University Medical Center, Leiden University, GGZ Rivierduinen, University Medical Center Groningen, University of Groningen, Lentis, GGZ Friesland, GGZ Drenthe, Rob Giel Onderzoekscentrum).

**PROspective Study of Pravastatin in the Elderly at Risk (PROSPER)**

The PROSPER study was supported by an investigator initiated grant obtained from Bristol-Myers Squibb. Prof. Dr. J. W. Jukema is an Established Clinical Investigator of the Netherlands Heart Foundation (grant 2001 D 032). PROSPER was supported by the European Federation of Pharmaceutical Industries Associations (EFPIA), Innovative Medicines Initiative Joint undertaking, European Medical Information Framework (EMIF) grant number 115372 and the European Commission under the Health Cooperation Work Programme of the 7th Framework Programme (Grant number 305507) “Heart ‘omics’ in AGEing” (HOMAGE).

**The LUMC arthroplasty studies**

This was a combination of TACTICS, TOMAAT and RAAK cohorts. TACTICS was funded by The Dutch Board of Health Care Insurances (College voor Zorgverzekeringen; OG99/023) and Sanquin Blood Bank. Involved were Prof. dr R.G.H.H. Nelissen, MD, Prof. dr A. Brand, MD, Leiden University Medical Centre; R.L. te Slaa MD, Reinier de Graaf Gasthuis, Delft; Dr R.G. Poll MD, Slotervaart ziekenhuis, Amsterdam; Dr K.M. Veenstra Franciscus ziekenhuis, Rotterdam and Prof. dr D. van Rhenen Sanquin Blood Bank, Rotterdam. Funding for the TOMAAT-study was received from ZonMW (06-601) and Sanquin Blood Supply (03-002), the Netherlands. Clinical Trial Number: ISRCTN96327523 (controlled-trials.com) and NTR 303 (Dutch Trial Register). The RAAK study was supported by the Leiden University Medical Centre. Furthermore, the molecular studies performed within to the RAAK study has received funding from the Dutch Arthritis Association (DAA_10_1-402), Biobanking and BioMolecular resources Research Infrastructure The Netherlands (BBMRI-NL) complementation project CP2013-84-CP2013-83 and Dutch Scientific Research council NWO /ZonMW VICI scheme (nr. 91816631/528).

**STEMI_GIPS-III**

The study received financial support from the Netherlands Organization for Medical Research (ZonMw; grant nr. 95103007); the funding source had no role in the study. GIPS-III 2010B257 The data are available upon request to all interested researchers provided their research question is within the informed consent provided by the participants. Patients did not provide informed consent to publicly release their data on an individual level on the internet. Data can be made available to other researchers upon request to Prof. Van der Harst after submitting a research proposal and approval of the GIPSIII steering committee responsible to ensure research question falls within the limits of the informed consent and IRB approval. The data will be released under a MTA to ensure future compliance to the obtained informed consent by other researchers.

**UCORBIO**

UCORBIO is conducted and supported by department of Cardiology, University Medical Center Utrecht, Netherlands. Folkert W. Asselbergs is supported by UCL Hospitals NIHR Biomedical Research Centre. Metabolic profiling was supported by Biobanking and BioMolecular resources Research Infrastructure, the Netherlands (BBMRI-NL). UCORBIO received funding from FP EU project CVgenes@target (HEALTH-F2-2013–601456). We would like to thank Ms. Jonne Hos and Ms. Merel Schurink for their logistical support and Daniel Kofink, PhD, for data management.

**VUNTR**

Funding was obtained from the Netherlands Organization for Scientific Research (NWO) and MagW/ZonMW grants 904-61-090, 985-10-002, 904-61-193,480-04-004, 400-05-717, Addiction-31160008, Middelgroot-911-09-032, Spinozapremie 56-464-14192, Biobanking and Biomolecular Resources Research Infrastructure (BBMRI –NL, 184.021.007).; the European Community's Seventh Framework Program (FP7/2007-2013), ENGAGE (HEALTH-F4-2007-201413); the European Science Council (ERC Advanced, 230374), Rutgers University Cell and DNA Repository (NIMH U24 MH068457-06), the Avera Institute, Sioux Falls, South Dakota (USA) and the National Institutes of Health (NIH, R01D0042157-01A, MH081802, Grand Opportunity grants 1RC2 MH089951). We gratefully acknowledge grant NWO 480-15-001/674: Netherlands Twin Registry Repository: researching the interplay between genome and environment.

**HOF**

Main funding for initial data collection and analysis for the HOF study was provided by U.S National Institutes of Health grant R01-HL067914 and for subsequent analyses by grants R01-AG042190 and R01-AG066887.

**STABILITEIT**

#### Ethics statements

**Alpha Omega Cohort**

The protocol of the *Alpha Omega Trial*, from which the *Alpha Omega Cohort* emerged, was approved by the Medical Ethics Review Committee of Haga Hospital "Leyenburg", The Hague (METC Zuidwest Holland; L01.049) and by local ethics committees of all participating hospitals. In accordance with the Declaration of Helsinki, the *Alpha Omega Cohort* obtained informed consent from all participants prior to their entering the study.

**Amsterdam Dementia Cohort**

The Amsterdam Dementia Cohort study protocol was approved by the Medical Ethics Committee of VU University Medical Centre (2016.061). In accordance with the Declaration of Helsinki, the Amsterdam Dementia Cohort obtained informed consent from all participants prior to their entering the study.

**BIOMARCS**

The *BIOMarker study to identify the Acute risk of a Coronary Syndrome (BIOMArCS)* study protocol was approved by the *Medical Ethics Committee of the Erasmus MC (MEC-2007-185)*. In accordance with the Declaration of Helsinki, the *BIOMarker study to identify the Acute risk of a Coronary Syndrome (BIOMArCS)* obtained informed consent from all participants prior to their entering the study.

**LUMINA**

The study protocols making up the *Leiden University MIgraine Neuro-Analysis* (LUMINA) cohort were approved by the Medisch Ethische Toetsingcommissie (METC) of the Leiden University Medical Center (LUMC) (registration numbers P07.079 and P12.113). In accordance with the Declaration of Helsinki, the LUMINA CSF, MRS, and CHARM subcohorts obtained informed consent from all participants prior to their entering the study.

**CHECK**

Cohort Heup en Cohort Knie study protocol was approved by Medical ethics committees of all participating centres. In accordance with the Declaration of Helsinki, the Cohort Heup en Cohort Knie obtained informed consent from all participants prior to their entering the study.

**CODAM**

The Cohort on Diabetes and Atherosclerosis Maastricht (CODAM) study protocol was approved by the Medical Ethics Review Committee of the AzM/UM (MEC 99-112 /MEC05-170). In accordance with the Declaration of Helsinki, the CODAM study btained informed consent from all participants prior to their entering the study

**The Maastricht Study (TMS)**

*The Maastricht Study* protocol was approved by the Medical Ethics Review Committee AzM/UM (NL31329.068.10) and the Minister of Health, Welfare and Sports of the Netherlands (Permit 131088-105234-PG). In accordance with the Declaration of Helsinki, the *Maastricht Study* obtained informed consent from all participants prior to their entering the study.

**The Hoorn Diabetes Care System cohort study (DCS)**

The Hoorn Diabetes Care System Cohort study protocol was approved by the ethical committee VU Vrije Universiteit Medical Center (2007/57). In accordance with the Declaration of Helsinki, the The Hoorn Diabetes Care System Cohort obtained informed consent from all participants prior to their entering the study.

**Erasmus Rucphen Family (ERF) study**

The Erasmus Rucphen Family study protocol was approved by the Medical Ethics Committee of the Erasmus MC Rotterdam, the Netherlands (MEC 213.575/2002/114). In accordance with the Declaration of Helsinki, the Erasmus Rucphen Family study obtained informed consent from all participants prior to their entering the study.

**Rotterdam Study**

*The Rotterdam S*tudy protocol was approved by the Medical Ethics Committee of the Erasmus MC Rotterdam, the Nethrlands. (*MEC 02.1015*) and by the Dutch Ministry of Health, Welfare, and Sport (Population Screening Act WBO, license number 1071272-159521-PG). In accordance with the Declaration of Helsinki, the *Rotterdam Study* obtained written informed consent from all participants prior to their entering the study.

**FUNCTGENOMICS**

*The 500 Functional Genomics* study protocol was approved by Commissie Mensgebonden Onderzoek Regio Arnhem-Nijmegen (42561.091.12). In accordance with the Declaration of Helsinki, the *500 Functional Genomics* study obtained informed consent from all participants prior to their entering the study.

**GARP**

The Genetics, ARthrosis and Progression study protocol was approved by the Medical Ethic Committee of the LUMC (P 76/98). In accordance with the Declaration of Helsinki, the Genetics, ARthrosis and Progression obtained informed consent from all participants prior to their entering the study.

**HELIUS**

*The HEalthy LIfe in an Uran Society (HELIUS)* study protocol was approved by the Medical Ethics Board of the Amsterdam University Medical Centers, location AMC (*METC 10/100#10.17.1729*). In accordance with the Declaration of Helsinki, the *HEalthy LIfe in an Uran Society study* obtained informed consent from all participants prior to their entering the study.

**LIFELINES-DEEP**

The LifeLines DEEP study was approved by the ethics committee of the University Medical Center Groningen, document number METC UMCG LLDEEP: M12.113965. All participants signed an informed consent form before study enrollment. All procedures performed in studies involving human participants were in accordance with the ethical standards of the institutional and/or national research committee and with the 1964 Helsinki declaration and its later amendments or comparable ethical standards.

**Leiden Longevity Study (LLS)**

The Leiden Longevity Study protocol was approved by the Medical Ethical Committee of the Leiden University Medical Center before the start of the study (P01.113). In accordance with the Declaration of Helsinki, the Leiden Longevity Study obtained informed consent from all participants prior to their entering the study.

**NESDA**

The Netherlands Study of Depression and Anxiety (NESDA) was approved by the Medical Ethical Committee of the VUmc (reference number 2003/183). In accordance with the Declaration of Helsinki, the Netherlands Study of Depression and Anxiety obtained informed consent from all participants prior to their entering the study.

**PROspective Study of Pravastatin in the Elderly at Risk (PROSPER)**

*The Prospective* study of Pravastatin in the Elderly at Risk protocol was approved by institutional ethics review boards of centres of Cork University (Ireland), Glasgow University (Scotland) and Leiden University Medical Center (the Netherlands). In accordance with the Declaration of Helsinki, the *The Prospective study of Pravastatin in the Elderly at Risk* obtained informed consent from all participants prior to their entering the study.

**The LUMC arthroplasty studies**

**RAAK**

Research Artrotisch Articulair Kraakbeen study protocol was approved by Medical Ethics Committee of the LUMC (P08.239 and P19.013). In accordance with the Declaration of Helsinki, the Research Artrotisch Articulair Kraakbeen obtained informed consent from all participants prior to their entering the study.

**TACTICS**

The 'Kwaliteit van leven en morbiditeit 10 jaar na totale heup of knie vervanging' study protocol was approved by Medical Ethics Committee of the LUMC (P00.179 and P11.050). In accordance with the Declaration of Helsinki, the Kwaliteit van leven en morbiditeit 10 jaar na totale heup of knie vervanging obtained informed consent from all participants prior to their entering the study.

**TOMAAT**

*The 'Transfusie op maat studie - optimale bloedmanagement bij electieve orthopedische ingrepen'* study protocol was approved by Medical Ethics Committee of the LUMC (P03.044). In accordance with the Declaration of Helsinki, the Transfusie op maat studie - optimale bloedmanagement bij electieve orthopedische ingrepen obtained informed consent from all participants prior to their entering the study.

**STEMI_GIPS-III**

The Glycometabolic Intervention as Adjunct to Primary Percutaneous Coronary Intervention in ST- Segment Elevation Myocardial Infarction (GIPS) III study is a double-blind placebo-controlled randomized clinical trial and was designed to determine whether metformin preserves left ventricular function after ST-segment elevation myocardial infaction (STEMI) in patients without diabetes. The study protocol was in accordance with the Declaration of Helsinki and was approved by the institutional review board (METC 2010.077, Groningen, the Netherlands) and national regulatory authorities. This trial was registered at clinicaltrials.gov (NCT01217307).

**UCORBIO**

Utrecht Coronary Biobank Study (UCORBIO) study protocol was approved by the Medical Ethics Committee of the UMC Utrecht (reference number 11–183). In accordance with the Declaration of Helsinki, the Utrecht Coronary Biobank Study (UCORBIO) obtained informed consent from all participants prior to their entering the study.

**VUNTR**

The Netherlands twin Register study protocol was approved by the Central Ethics Committee on Research Involving Human Subjects of the VU University Medical Centre, Amsterdam, an Institutional Review Board certified by the U.S. Office of Human Research Protections (IRB number IRB00002991 under Federal-wide Assurance-FWA00017598. In accordance with the Declaration of Helsinki, the Netherlands Twin Register obtained informed consent from all participants prior to their entering the study.

**HOF**

The HOF study “Prenatal nutrition and adult disease, an analysis of sib-pairs discordant for exposure to the Dutch famine of 1944-1945” was approved by the Institutional Review Board of Columbia University, New York (IRB-AAAB4053) and by the Medical Ethics Committee of Leiden University Medical Center (P02.082). Written informed consent was obtained from all study participants.

**STABILITEIT**

The Amsterdam Dementia Cohort study protocol, from which the STABILITEIT cohort emerged, was approved by the Medical Ethics Committee of VU University Medical Centre (2016.061). In accordance with the Declaration of Helsinki, the Amsterdam Dementia Cohort obtained informed consent from all participants prior to their entering the study.
